# Variant-to-function mapping in lupus links *IL12A* to the expansion of disease-associated B cells with a cytotoxic program

**DOI:** 10.64898/2026.07.28.26359131

**Authors:** Mariasilvia Colantuoni, Qian Xiao, Nada Abdel Aziz, Vitor R. C. Aguiar, Sarah Djeddi, Yifei Liao, Daniela Fernandez-Salinas, Taehyeung Kim, Laura B. Lewandowski, Jinghua Gu, Uthra Balaji, Tracey Wright, Joyce C. Chang, Virginia Pascual, Peter A. Nigrovic, Benjamin E. Gewurz, Maria Gutierrez-Arcelus

## Abstract

Systemic lupus erythematosus (SLE) is a chronic autoimmune disease characterized by over 200 risk variants identified through genome-wide association studies. While the majority of these are non-coding variants with unresolved functions, elucidating their mechanisms is critical for prioritizing therapeutic targets with increased clinical success. Several SLE risk loci span genes involved in the IL-12 signaling pathway. However, for most of them the causal variants, their definitive target genes, and their cellular consequences remain unestablished. Concurrently the expansion of double-negative 2 (DN2) B cells is a hallmark of SLE, but whether IL-12 and/or genetic risk functionally drive DN2 cells is unclear. In this study, we integrated candidate risk variants at the 3q25.33 risk locus with regulatory maps of a B cell line, identifying risk variant rs485499 located within a putative enhancer 39kb downstream of *IL12A*, and overlapping an open chromatin region in primary B cells stimulated with a DN2-skewing cocktail. Using CRISPR-based tools in a B cell line, we validated this region as an enhancer, rs485499 as a likely causal variant and established *IL12A* as its definitive target gene. Individuals homozygous for the rs485499 risk allele exhibited elevated *IL12A* production in naïve B cells and presented with an expanded DN2 population in peripheral blood, compared to non-risk allele carriers. Mechanistically, we found the transcription factor IRF4 preferentially binds the rs485499 risk allele, driving *IL12A* upregulation. *In vitro* recombinant IL-12A promoted DN2 differentiation an effect that is abrogated by IL-12 inhibition with ustekinumab, establishing a causal IL-12–driven DN2 B cell expansion axis. Finally, we reveal that DN2 B cells inherently possess a previously unrecognized cytotoxic function that is potentiated by the IL-12 signaling axis. This cytotoxic profile is further supported by SLE patient data, identifying it as a bona fide effector state of DN2 B cells. Collectively, these findings identify rs485499 as a likely causal variant within this locus and establish a functional link between genetic risk and DN2 B cell expansion, validating this subset as a key pathogenic driver armed with a newly identified cytotoxic program. Furthermore, we identify the IL-12–IFNy–DN2 axis as a promising therapeutic target, providing a mechanistic rationale for future subset-specific interventions in SLE.

## INTRODUCTION

Systemic lupus erythematosus (SLE) is a debilitating autoimmune disorder predominantly affecting women, ranking among the leading causes of death in young females^1,2^. Characterized by the production of autoantibodies and multi-organ inflammation, SLE arises from a complex interplay between genetic predispositions and environmental factors^3^. Genome-wide association studies (GWAS) have identified more than 200 risk loci for SLE, the vast majority of which implicate non-coding variants^4–10^. Some of these non-coding risk variants are enriched in regulatory elements specific to B cells^11,12^. SLE risk variants are particularly enriched in open chromatin regions of B cells activated by TLR7 and BCR agonists, in conjunction with IFN-γ, IL-21, IL-2, and BAFF^13^, which together can induce the differentiation of naïve B cells into DN2 (IgD⁻ CD27⁻ CD21⁻ CD11c⁺) B cells, a relevant subset in autoimmunity^14–16^. This suggests that an important proportion of SLE risk loci influence gene regulation in B cells on their way to DN2 B cell differentiation. However, this supposition remains to be established.

DN2 B cells, also known as age- or autoimmune-associated B cells (ABCs) especially in mouse studies, are a unique subset of B cells that in mice increase with age and drive autoimmunity^17,18^. In humans, DN2 B cells are expanded in multiple autoimmune conditions, such as SLE^14,19–21^, rheumatoid arthritis^22^, multiple sclerosis^23^, autoimmune anemia^24^, scleroderma^14^, Sjögren’s syndrome^14^, and Crohn’s disease^25^. Most typically, human DN2 B cells are defined as either CD19+ IgD⁻ CD27⁻ CD11c⁺ B cells or CD21⁻ CD11c⁺ B cells. DN2 exhibit a shared clonality with plasma cells and express elevated levels of IRF4, a transcription factor critical for plasma cell differentiation^26–29^, and its target genes^14,19,30^. A probable mechanism by which DN2 B cells contribute to autoimmunity is through the generation of autoantibodies, as supported by mouse models and observations in humans^18,31,32^. In SLE patients, the abundance of DN2 B cells correlates with autoantibody levels^14,32,33^. These observations identify the DN2 B cell phenotype as a pre-plasma cell state poised for rapid antibody production and support a likely pathogenic role in SLE^14,19,31^.

Although the full spectrum of signals driving DN2 B-cell differentiation is still being defined, it is well established that DN2 cells arise in an environment rich in IFN-γ, which acts as a primary driver of their development^14,15^. IFN-γ, together with IL-21, synergizes with BCR and TLR7 engagement to push naïve B cells toward the DN2 phenotype. Recently, it has been shown that IL-12 promotes plasma cell differentiation via an IL-12/IFN-γ feed-forward loop^34,35^, raising the possibility that IL-12 could also have a role in DN2 B cell differentiation.

Notably, part of the genetic susceptibility to SLE is strongly linked to the IL-12 signaling pathway. GWAS have identified risk variants within loci encompassing *IL12A*, *IL12RB2*, *STAT4*, *IL12B*, and *TYK2*^6,7,9^. Within the 3q25.33 locus specifically, which contains the *IL12A* gene, two GWAS have identified a genome-wide significant risk signal^6,7^, although the precise target gene has not yet been determined. This locus is also associated with several other autoimmune diseases, including primary biliary cirrhosis (PBC)^36^, multiple sclerosis^37^, scleroderma^38^, celiac disease^39^, and Sjögren’s syndrome^40^.

In this study, we identify a likely causal SLE variant at 3q25.33 linked to altered *IL12A* expression through impaired IRF4 binding, which is associated with expansion of DN2 B cells. Using a combination of *in vitro* approaches and primary patient sample validation, we report that DN2 B cells exhibit cytotoxic features. These findings reveal a genetic pathway by which SLE genetic risk acts through the IL-12/IFN-γ axis to promote DN2 B cell expansion, offering mechanistic evidence for the pathogenicity of DN2 B cells in SLE. Collectively, our results provide new insights into the genetic mechanisms driving SLE and highlight the IL-12/DN2 axis as a potential target for therapeutic intervention.

## RESULTS

### *In silico* identification of candidate regulatory SLE risk variants at the 3q25.33 locus in B cells

SLE GWAS have identified risk variants within the 3q25.33 locus, which harbors many single-nucleotide polymorphisms (SNPs) in high linkage disequilibrium (LD) with the lead variant rs564976^7^ (**Fig. 1a**). Although *IL12A* is the nearest gene, its role as the target gene remains uncertain. We leveraged public expression quantitative trait locus (eQTL) data from multiple tissues and cell types^41^ to assess the association between genetic variation and gene expression levels. Colocalization analysis between the SLE risk locus and *IL12A* eQTLs from Epstein–Barr virus (EBV) transformed B cells (lymphoblastoid cell lines, LCLs) from the TwinsUK^42^ and Geuvadis^43^ cohorts revealed the highest posterior inclusion probabilities (PIP) for the hypothesis that there is a shared causal variant between the risk locus and the eQTL (PP4 = 0.953 and 0.94, respectively) (**Extended Table 1**). These results suggest that a genetic variant at 3q25.33 may increase SLE risk by modulating *IL12A* expression levels within the B cell lineage. Given the eQTL evidence in B cell lines, the relative underrepresentation of B cells in genetic studies, and the well-established role of B cells in SLE^44–46^, we chose to carry out further analyses in the GM12878 LCL model. A primary advantage of the GM12878 model is its status as a Tier 1 ENCODE cell line, providing access to a vast repository of public epigenetic and functional genomic data^47^. We sought to identify candidate causal variants in the locus. First, we prioritized SNPs within 3q25.33 with *p* = 1e-5 and in high linkage disequilibrium with the lead variant (r² > 0.8) based on data from Langefeld et al., GWAS^7^ (**Methods).** This approach yielded a candidate set of 63 proxy variants (*n* = 63) (**Extended Table 2**). Next, we assessed the overlap of these variants with predicted-enhancer regions by ChromHMM which used GM12878 chromatin marks by ENCODE^47^, identifying 36 putative enhancer-associated variants located within active or poised regulatory elements. We further verified the intersection of these variants with GM12878 ENCODE chromatin marks^47^ and GM12878 nascent transcripts (that can tag active enhancer RNAs)^48^. These features included DNase I hypersensitivity regions (DNase-seq), ChIP-seq peaks for H3K4me1, H3K4me3, and H3K27ac, open chromatin regions by ATAC-seq, and nascent transcripts by GRO-seq (**Fig. 1a and Extended Table 2**). Through this integrative approach, rs485499, located 39Kb downstream of *IL12A*, emerged as the strongest candidate regulatory variant, overlapping the greatest number of regulatory features in GM12878 (**Fig. 1b**). In addition, using our ATAC-seq data in primary B cells^13^, we observed rs485499 overlaps an open chromatin region specific to B cells stimulated with the DN2-inducing cocktail, compared to non-stimulated B cells or B cells activated with other stimuli (**Fig.1c**). Furthermore, rs485499 has the highest PIP for *IL12A* expression in a fine-mapping multi-ancestry LCL eQTL study^49^ and it is the lead variant in the 3q25.33 locus in a recent SLE-enriched multi-ancestry multi-trait genome-wide association meta-analysis^50^. Together, these analyses point to rs485499 as a candidate causal variant in the 3q25.33 SLE risk locus that could be altering *IL12A* gene regulation in B cells.

**Figure 1.**
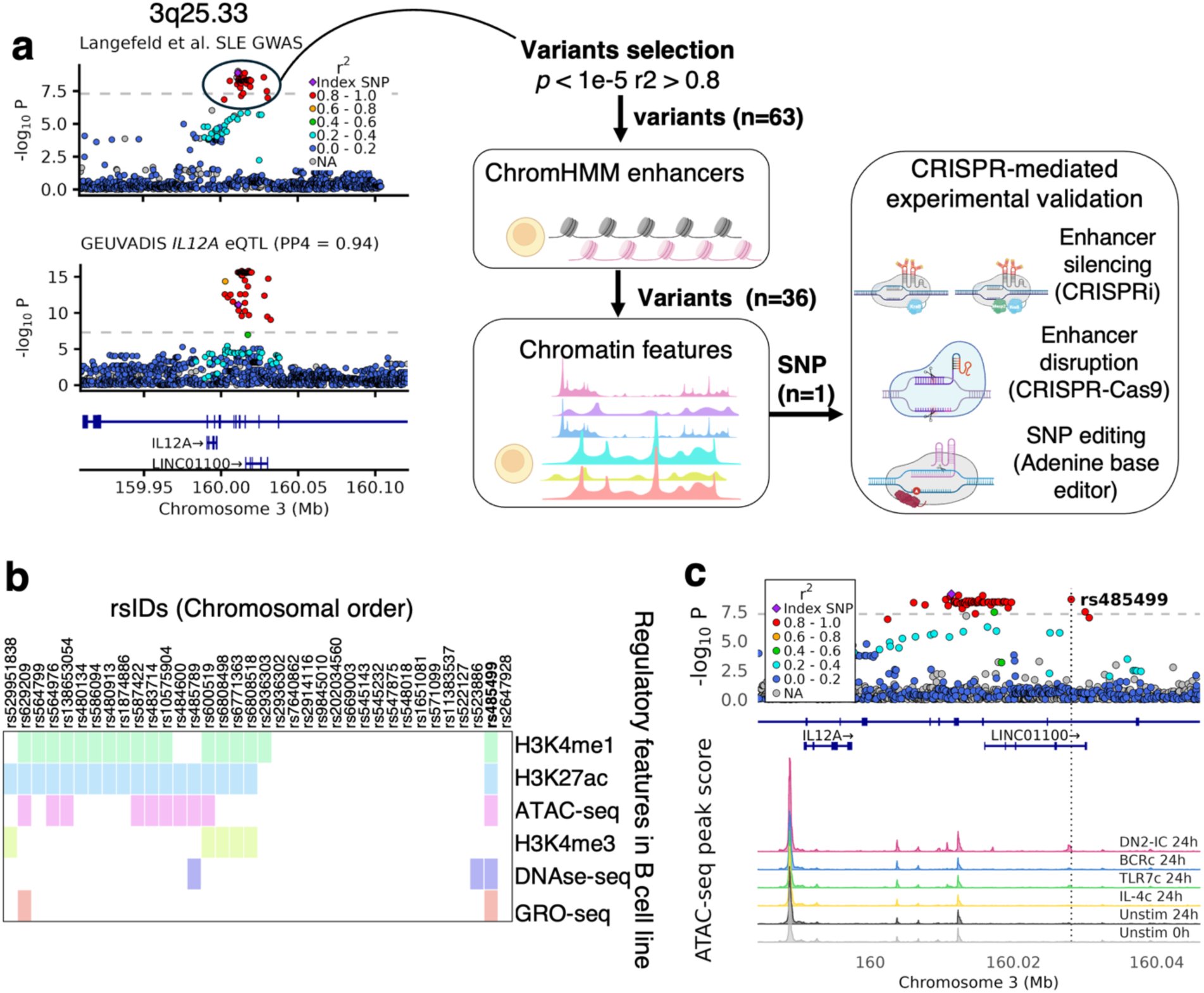
In silico and experimental workflow to identify rs485499 as a candidate causal variant at the 3q25.33 SLE risk locus in B cells. **(a) Left panel.** Colocalization between SLE GWAS 3q25.33 risk locus (top) and *IL12A* eQTL in LCLs (b= 0.94) (bottom). The purple diamond indicates the lead GWAS variant, while red dots indicate variants in high linkage disequilibrium (LD) with it. **Right panel**. Schematic representation of the workflow used to identify candidate causal variants from the 63 variants in high linkage disequilibrium with the lead variant at the 3q25.33 risk locus (selected from Langefeld et al. GWAS). The pipeline integrates ChromHMM-predicted enhancers and chromatin features to prioritize candidate regulatory variants for experimental validation using a modular CRISPR toolkit, including CRISPRi, CRISPR-Cas9, and base editing. **(b)** Heatmap of 36 variants within ChromHMM-predicted enhancer regions showing overlap (with colored boxes) with DNase I hypersensitivity regions (DNase-seq, ChIP-seq peaks for histone modifications (H3K4me1, H3K4me3, H3K27ac), open chromatin regions identified by ATAC-seq, and nascent transcripts, such as enhancer RNAs, identified by GRO-seq. Rs485499 is identified as the SNP with the highest number of regulatory chromatin hits. **(c)** Data from Aguiar et al. *medRxiv* showing the intersection of rs485499 with an open chromatin region specific to primary B cells stimulated with the DN2-IC (DN2 inducing cocktail). Top panel indicates the GWAS summary statistics from Langefeld et al. for the 3q25.33 locus. Primary B cells were cultured under the following experimental conditions: Unstim 0h: Unstimulated baseline control cells sampled at 0 hours. Unstim 24h: cells cultured in media alone for 24 hours. BCR 24h: Cells stimulated with anti-IgG/IgM antibodies and IL-4 for 24 hours. TLR7c 24h: Cells stimulated with the TLR7 agonist resiquimod (R848) for 24 hours. IL-4c 24h: Cells stimulated with IL-4 alone for 24 hours.

### CRISPR-based validation of rs485499 as a putative causal variant at the 3q25.33 locus

We next sought to experimentally test whether rs485499 represents a regulatory variant in the 3q25.33 locus and whether *IL12A* is the target gene using multiple independent CRISPR-based approaches (**Fig. 1a**). First, to functionally validate the putative enhancer overlapping rs485499, we generated a GM12878 cell line stably expressing dCas9 fused to the repressor domain KRAB for CRISPR interference (CRISPRi) epigenetic silencing, following previously published protocols^51^. We designed a single guide RNA (sgRNA) targeting the putative enhancer containing rs485499. Additionally, we included a sgRNA targeting the region harboring the lead GWAS variant rs564976. CRISPRi-mediated repression of the rs485499-containing region resulted in a marked reduction in *IL12A* expression, whereas targeting the region containing rs564976 had no significant effect (**Fig. 2a**). We validated the regulatory activity of the rs485499-containing region on the *IL12A* gene through two additional independent, complementary methods. Targeted inactivation via dCas9-KRAB-MeCP2 (a CRISPRi method that uses two repressor domains) (**Fig. 2b**) confirmed the enhancer’s role in transcriptional control of *IL12A*, a finding reinforced by the subsequent CRISPR-Cas9-induced disruption (CRISPR-Cas9) of the region (**Fig. 2c**). These results indicate that rs485499 resides within an enhancer regulating *IL12A* expression.

**Figure 2.**
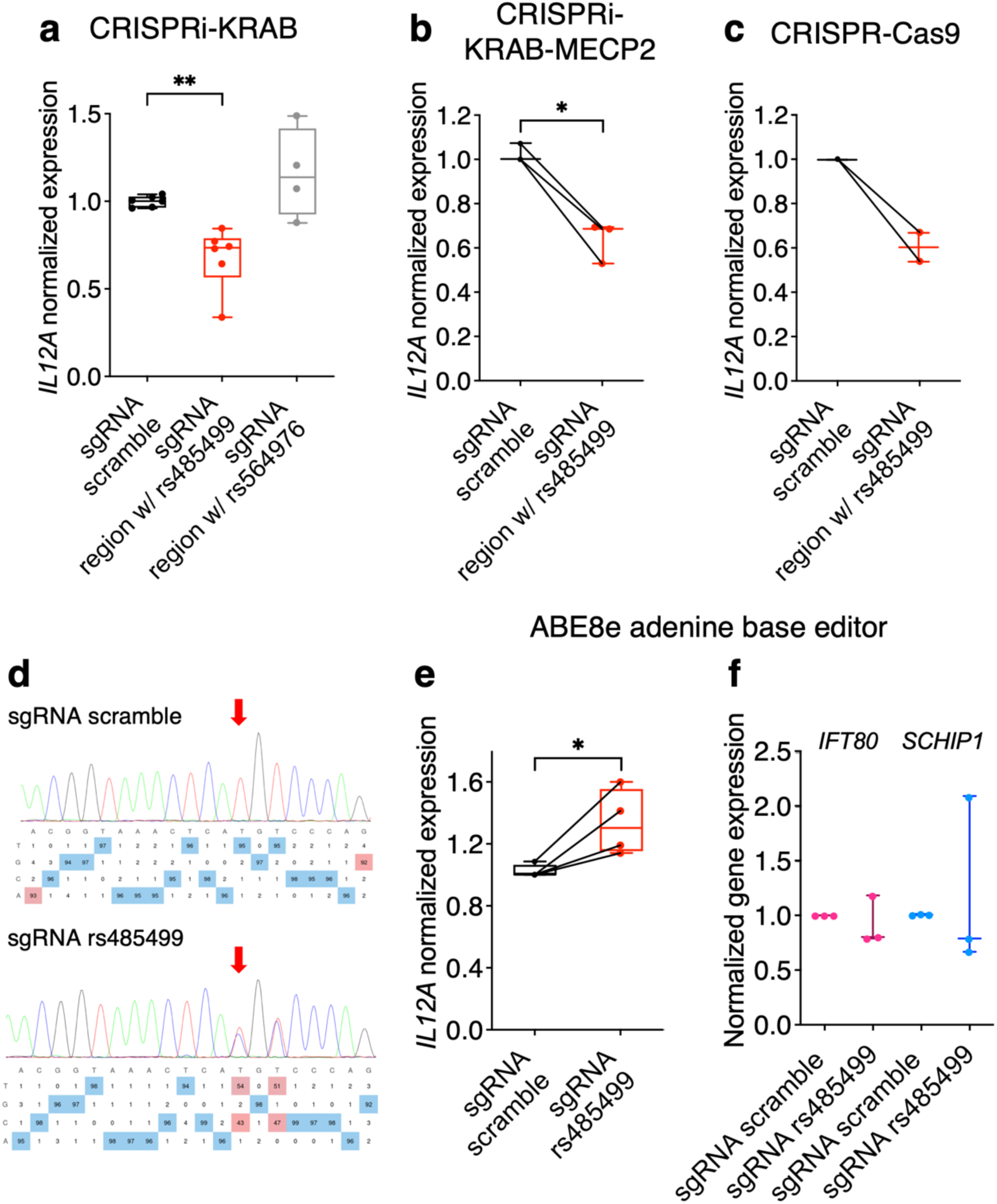
CRISPR-based validation of rs485499 as a likely causal variant at the 3q25.33 risk locus. **(a)** Enhancer silencing with dCas9-KRAB based CRISPRi of the region containing rs485499 (*n* = 6) and the GWAS lead variant rs564976 (*n* = 4). **(b)** Validation with dCas9-KRAB-MECP2-CRISPRi and **(c)** CRISPR Cas9 knock-out of the region containing rs485499. **(d)** Adenine base editing of rs485499 (resulting in a genomic T-to-C transition), where red arrows indicate the specific change T to C at rs485499, and **(e)** shows the resulting *IL12A* expression change (*n* = 4). **(f)** Gene expression changes observed for neighboring genes *IFT80* and *SCHIP1* at the 3q25.33 locus (*n* = 3). Data are presented as box plots showing the median and 25th/75^th^ percentiles; whiskers indicate the range, and individual data points represent biological replicates (*n* = 3 to 6). Data from two independent experimental runs were combined and plotted together in panels **a** and **e**. Two-tailed paired t test. \**p* < 0.05, \*\**p* < 0.01. Gene expression is normalized to *GAPDH* and expressed relative to the non-targeting scramble control using the ΔΔCt method.

To further assess whether rs485499 itself regulates *IL12A*, we employed a base editing approach in GM12878 cells. Because GM12878 cells are homozygous for the rs485499 risk allele (T), we generated a cell line stably expressing an adenine base editor (ABE8e) fused to dCas9, using lentiviral particles as in our CRISPRi experiments^51^. We designed a sgRNA to convert the rs485499 T allele (targeting the complementary A on the opposite strand) to the non-risk C allele. Base editing achieved approximately 50% efficiency at rs485499; however, it also edits an adjacent A (position 4 of the sgRNA) (**Fig. 2d**). Following nucleofection of GM12878 cells with the sgRNA, we observed an increase in *IL12A* expression levels, consistent with eQTL data in LCLs, where the risk allele is associated with decreased *IL12A* expression^42,43^ (**Fig. 2e**).

Given that enhancer–promoter interactions often occur within 200-300kb^52^, we selected a window spanning 500 kb (250 kb upstream and downstream of the lead GWAS variant rs564976) and identified three additional genes in the locus: *IFT80*, *C3ORF80*, and *SCHIP1*. We then examined gene expression changes upon base editing and found no detectable effect for *IFT80* and *SCHIP1* (**Fig. 2f**). *C3ORF80* was not expressed in GM12878 cells. Collectively, these findings indicate that rs485499 is a likely causal variant within the 3q25.33 SLE risk locus, regulating *IL12A* expression in a B cell line.

### IL-12A promotes DN2 B cell differentiation through activation of the IL-12 signaling pathway

Transcriptomic analyses identify B cells as major producers of *IL12A* transcripts among immune cells^53,54^. However, IL-12A protein (p35) alone is poorly secreted and requires pairing with either IL-12B (p40) or EBI3 to form the functional cytokines IL-12 and IL-35, respectively^55–58^. Given that IL-12A is the rate-limiting subunit for the assembly of the L-12 heterodimer (p70)^59^, understanding its regulation in B cells, particularly within naïve subsets that maintain basal steady-state expression, is critical. On the contrary, IL-12 expression is well-characterized as being robustly inducible upon stimulation with specific inflammatory signals, such as TLR ligands, CD40L, and IFN-y ^60–62^. In our human B cell transcriptomic resource^13^, *IL12A* is upregulated early after B cell activation with a DN2-inducing cocktail (DN2-IC) **(Extended Fig.1a**) designed to mimic the SLE microenvironment and that contains anti-IgG/IgM/IgA (to engage the BCR), resiquimod (R848) to activate TLR7, B cell activating factor (BAFF), IL-2, IL-21, and IFN-γ, as previously reported^14,15,63^. We confirmed this expression profile with qPCR in naïve B cells activated with the DN2-IC, showing upregulation at 8 hours followed by decline in expression (**Fig. 3a**). We confirmed in vitro naïve B cell differentiation into CD19⁺ IgD⁻ CD27⁻ CD21⁻ CD11c⁺ DN2 B cells with the DN2-IC using flow cytometry (**Fig. 3b**). To investigate the biological role of *IL12A* during naive B cell differentiation into DN2 B cells, we employed two complementary experimental approaches (**Fig. 3c**). First, we performed a CRISPR-mediated knockout of *IL12A* in naïve B cells undergoing DN2 differentiation in vitro, using 4 to 6 donors. One hour after DN2 B cell stimulation, Cas9 protein and a pool of three sgRNAs targeting *IL12A* exon 2 were introduced by nucleofection. We estimated an editing efficiency of approximately 50% at 48 hours post-nucleofection (**Extended Fig. 1b**). *IL12A* knockout resulted in reduced frequencies of DN2 B cells at days 3 and 6 post-nucleofection (*p* = 0.0259 and *p* = 0.0069) (**Fig. 3d**) compared to the scramble sgRNA control. However, because *IL12A* is upregulated so early after stimulation (**Fig. 3a**), it is likely that gene disruption occurred after the initial transcriptional burst, which may explain the modest impact on early DN2 differentiation. Conversely, DN2-IC supplementation with recombinant human IL-12A (rhIL-12A) markedly enhanced DN2 differentiation at both day 3 and day 6 (*p* = 0.02 and *p* = 0.0003 **Fig. 3d**) compared to activation with DN2-IC alone (**Fig. 3d**), with total CD11c-expressing B cells following similar dynamics (**Extended Fig. 1c**). Both perturbations did not significantly alter the overall DN population (CD19+ IgD⁻ CD27⁻) (**Fig. 3e**) and the other minor populations obtained in culture (**Extended Fig. 1d**). Furthermore, we assessed the expression of two key DN2 B cell markers, T-bet and IFN-γ, both of which were significantly upregulated following IL-12A treatment during differentiation (**Extended Fig. 1e**). Together, these results show that IL-12A promotes DN2 B cell differentiation.

**Figure 3.**
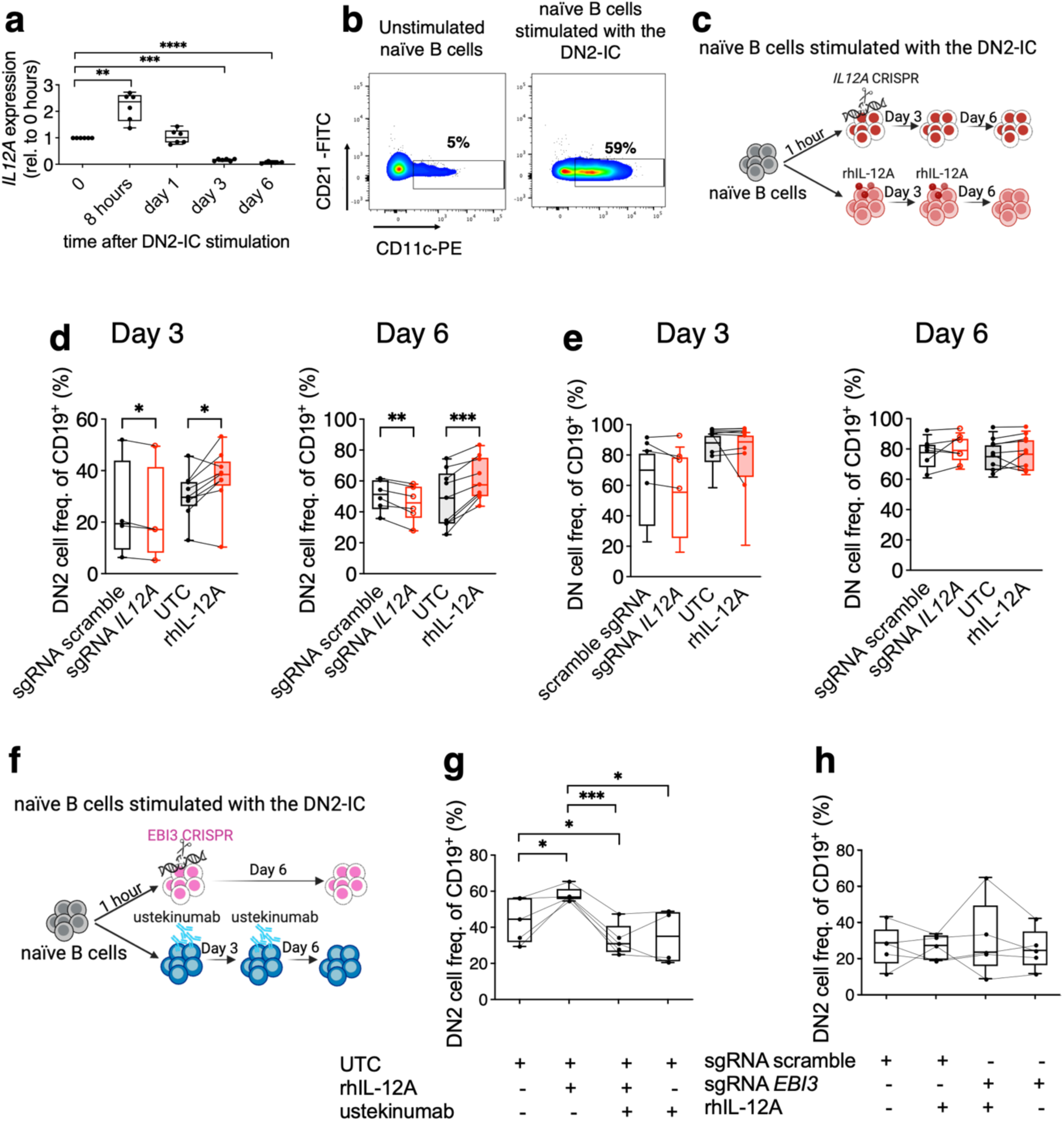
IL-12A promotes DN2 B cell differentiation through IL-12 signaling. **(a)** *IL12A* expression kinetics in naïve B cells stimulated with DN2-IC. **(b)** Representative FACS plots illustrating the expansion of DN2 B cells (CD19^+^ IgD⁻ CD27⁻ CD21⁻ CD11c⁺) at day 6 following in vitro naïve B cell differentiation with DN2-IC. **(c)** Experimental schematic for *IL12A* KO or recombinant IL-12A treatment during DN2-IC stimulation of naïve B cells. **(d–e)** Results for DN2 cell **(d)** and DN cell **(e)** differentiation following *IL12A* KO or recombinant IL-12A supplementation at days 3 and 6. **(f)** Schematic of *EBI3* KO or ustekinumab (anti-IL-12B) treatment during DN2-IC stimulation. Effect of ustekinumab **(g)** and *EBI3* KO **(h)** on IL-12A-dependent DN2 B cell differentiation. Data are presented as box plots showing the median and 25th/75th percentiles; whiskers indicate the range, and individual data points represent biological replicates (donors, *n* = 4 to 9). Data from 3-4 independent experimental runs were combined and plotted together in panels **d** and **e**. Cell populations are calculated as a percentage of total CD19^+^ B cells. Scramble sgRNA: cells nucleofected with the scramble sgRNA. SgRNA *IL12A*/sgRNA *EBI3*: cells nucleofected with sgRNA targeting *IL12A* or *EBI3* gene. UTC: untreated control, naive B cells stimulated with the DN2-IC. RhIL-12A: naive B cells stimulated with the DN2-IC in the presence of rhIL12A (200ng/ml). Ustekinumab: naive B cells stimulated with the DN2-IC in the presence of ustekinumab (10ug/ml). For all primary qPCR datasets, including the larger validation cohorts in Figure 6 showing *IL12A* expression over time during DN2-IC, *RPL10A* was used as the standard housekeeping control due to its superior stability during DN2-IC stimulation. In panel **(a)**, where *GAPDH* expression fluctuated during the initial differentiation time-course, gene expression is expressed relative to the Ct values at time 0 (naïve B cells). The expression trend was independently validated in separate experiments using *MTHFR* as a stable reference and in figure 6a, showing consistent results with the time 0 normalization. Two-tailed paired t test, \**p* < 0.05, \*\**p* < 0.01, \*\*\**p* < 0.001.

Both *IL12B* and *EBI3* were found to be upregulated during DN2 B cell differentiation (**Extended fig. 2a-b**). To determine whether IL-12 or IL-35 contributes to the IL-12A– driven DN2 expansion, we applied blocking strategies (**Fig. 3f**). We used ustekinumab, which neutralizes IL-12B, to inhibit IL-12 production, and because there is no commercially available blocker/inhibitor for EBI3, we CRISPR-knocked out *EBI3* (**Extended Fig. 2c**) to impede IL-35 formation. We observed a reduction in DN2 B cell generation at both day 3 and day 6 of differentiation in cultures treated with recombinant IL-12A and ustekinumab, compared with cultures treated with recombinant IL-12A alone (*p* = 0.0004, **Fig. 3g** and **Extended Fig. 2d**). In contrast, *EBI3* knockdown had no significant effect on DN2 differentiation (**Fig. 3h**). Together, these findings indicate that the IL-12A mediated promotion of DN2 differentiation depends on IL-12 signaling and is independent of IL-35.

### The IL-12 signaling axis potentiates a novel intrinsic cytotoxic program in DN2 B cells

To gain a deeper insight into IL-12A mediated DN2 expansion, we examined the longitudinal effect of IL-12A on the transcriptome for B cells undergoing DN2 differentiation. We performed single-cell RNA sequencing (scRNA-seq) on naïve B cells derived from five healthy donors, stimulated with the DN2-inducing cocktail in the presence (IL-12A stimulation) or absence (un-treated control, UTC) of rhIL-12A. Cells were profiled at day 1, 3, and 6 post-stimulation, totaling 50,883 cells post-QC (**Fig 4.a**) and visualized via uniform manifold approximation and projection (UMAP) by day and condition (**Fig. 4b-c**). Unsupervised clustering grouped cells into 5 main cell populations (**Fig. 4d-e and Extended Fig. 3a**): 1) *TBX21*^high^ (TBX21hi) activated B cells expressing genes associated with B cell activation such as *CD69* and high levels of IFN-response genes such as *IFI30* and *IFI35* which are primarily detected on day 1; 2) DN2-like cells expressing DN2-associated genes (*ITGAX*, *FCRL5*, *ZEB2*, *TNFRSF1B*) dominated day 6; 3) proliferative DN2-like cells co-expressing aforementioned DN2 markers and proliferative markers such as cell cycle-related genes (e.g. *MKI67*) and DNA-replication-associated genes (*TOP2A*, *PCNA* and *CDK1*), which are primarily found on day 3; 4) plasmablast-like and 5) plasma cell-like characterized by the expression of immunoglobulin genes and key transcription factors (e.g. *ATF5*, *XBP1*, *PRDM1*, *IRF4*), as well as some DN2 markers. Specifically, DN2-like and proliferative DN2-like clusters showed increased frequency on day 3 and day 6. In line with our FACS data, we showed that IL12A stimulation promoted the expansion of the DN2 subset, measured by an increase in the percentage of the DN2-like cluster in the IL12A stimulation condition (42%) compared to the UTC (34%) relative to the total cellular composition across timepoints (**Extended Fig. 3b**). As cell states are inherently continuous, we defined a transcriptional DN2 score derived from a bulk RNA-seq study on sorted B cell subsets from SLE patients and healthy controls^21^ (**Methods**). To further validate this DN2 identity scoring system, we applied it to an independent published scRNA-seq dataset^21^. We observed an overlap between the CellTypist reference-based automated annotations of ABCs^64^ and our continuous DN2 score (**Extended Fig. 3c-d**). Consistent with our flow cytometry observations and transcriptional cluster annotation, the continuous DN2 score captured the gradual increase of the DN2 state over time, reaching its maximum at day 6 (**Fig. 4f**). To investigate gene expression changes induced by IL-12A stimulation at each timepoint, we performed pseudo-bulk differential expression analysis between the IL-12A stimulation and UTC by day with linear mixed-effect models (LMM) adjusted for donor as a random effect (**Methods, Fig. 4g**). Consistent with our flow cytometry data, *IFNG* was among the top upregulated gene in IL-12A-stimulated cells on day 3 and day 6, confirming active IL-12 signaling (**Fig. 3 and Extended Fig. 1**). Furthermore, we observed significant upregulation of genes associated with IL-18 signaling (*IL18R1* and *IL18RAP*) and the granzyme family (*GZMB* and *GZMH*). Conversely, downregulated genes included the immunomodulatory factors *IDO1* and *IDO2*, as well as *AREG*. Gene Set Enrichment Analysis revealed a pattern of progressive transcriptional response to IL-12A treatment, with some pathways showing significant downregulation at day 1 and marked induction by days 3 and 6 (**Fig. 4h**). Notably we observed this trend in the Gene Ontology Biological Process leukocyte-mediated cytotoxicity (GO:0001909), with statistically significant enrichment at day 6 (*FDR* = 0.042) (**Extended Fig. 3e)**. The leading edge of the leukocyte-mediated cytotoxicity gene set included known cytotoxicity-related genes— such as *UNC13D*, *GZMB*, *HAVCR2*, and *NKG7*—whose expression increased by varying magnitudes on days 3 and 6 of IL-12A stimulation (**Extended Fig. 3f**). To quantify the overall upregulation of cytotoxicity at the transcriptional level, we designed a cytotoxicity score based on a curated gene set (*n* = 17) that includes cytotoxicity-related genes based on post-day 1 differentially expressed genes, leading edges in significant relevant GSEA pathways, and prior knowledge^65,66^ (**Methods).** We observed an overall progressive increase in cytotoxicity score over time, which as expected, was further elevated under IL-12A stimulation compared to UTC (**Fig. 4i**). In addition, per donor cytotoxicity score stratified by stimulation condition and colored by day showed strong positive correlation with DN2 score (rhIL-12A ρ = 0.886, UTC ρ = 0.964, **Fig. 4j**). Overall, these results suggest that naïve B cells differentiated *in vitro* toward the DN2 fate acquire a previously unrecognized cytotoxic transcriptional program, which is potentiated by IL-12 signaling.

**Figure 4.**
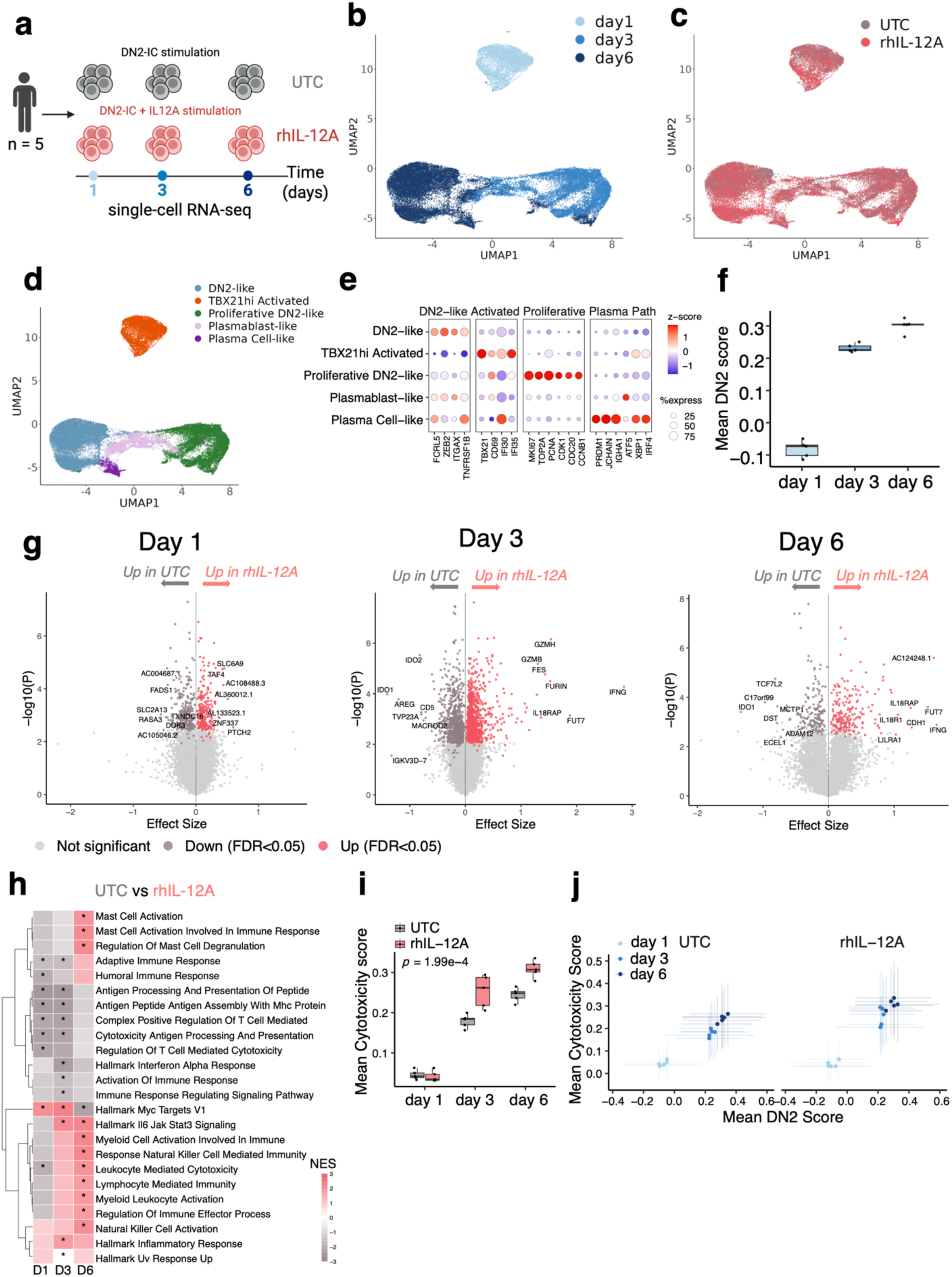
IL-12A drives a distinct transcriptional program during *in vitro* DN2 B cell differentiation. **(a)** Schematic representation showing RNA-seq profiling of Naïve B cells isolated from 5 donors stimulated with the DN2-IC in the absence (untreated control, UTC) or presence of rhIL-12A (IL-12Astimulation). Samples were harvested at day 1, 3, and 6 post-stimulation. **(b-d)** Uniform manifold approximation and projection (UMAP) visualization of 50,883 cells colored by **(b)** timepoint (day 1, 3, and 6), **(c)** stimulation condition (UTC vs. rhIL-12A) and **(d)** major cell types. **(e)** Dot plot showing expression of marker genes used for cell-state identification with z-score. **(f)** Box plot showing median DN2 score per donor across time points. **(g)** Volcano plots displaying the results of pseudobulk differentially expressed gene (DEG) analysis comparing IL-12A-stimulated samples to UTC controls at each individual time point. x-axis and y-axis respectively denote the effect size and log-transformed p-value from the linear mixed models. **(h)** Heatmap showing the normalized enrichment score (NES) of selective pathways from gene set enrichment analysis (GSEA). **(i)** Box plot showing mean cytotoxicity score per donor across timepoints, stratified by stimulation condition. P-value is obtained from linear-mixed model interaction term (cytotoxicity score x day) with sample as random effect. **(j)** Scatter plot showing mean cytotoxicity score vs. mean DN2 score per donor, stratified by stimulation condition and colored by timepoint. The error bars span the standard deviations within the donor. In (**h**), asterisks (*) denote adjusted *p-value* < 0.05.

To determine whether the cytotoxic signature observed in our *in vitro*-generated DN2 B cells could be captured in patients, we interrogated a published scRNA-seq dataset of peripheral blood mononuclear cells (PBMCs) derived from a cohort of children (cSLE) and adults with SLE (aSLE) (*n* = 41) and age-matched healthy controls (cHD and aHD, *n* = 17)^21^ (**Fig. 5a)**. We processed the sequencing data (**Extended Fig. 3c and methods**), extracted the B cells and plasma cells and calculated DN2 and cytotoxicity scores per cell (**Extended Fig. 3d and 3g and Methods**). Consistent with our *in vitro* findings, we observed a significant positive correlation between the DN2 score and cytotoxicity score across all donors (ρ = 0.36, *p* = 0.006, **Fig. 5b**). This pattern was accompanied by a significantly higher frequency of DN2-like B cells - defined by binarizing the DN2 score at an empirically determined threshold (**Extended Fig. 3h**) - in SLE compared to healthy donors (*p* = 0.006) consistent with the known expansion of DN2 B cells in SLE^14,19,20^ (**Fig. 5c**). Patients’ DN2-like B cells indeed preferentially expressed cytotoxicity-associated genes compared to other B cell populations (**Fig. 5d**). Within SLE patients, DN2-like cells showed significantly higher cytotoxicity scores than other B cells at different *IL12A* expression level (paired Wilcoxon, top 30%: *p* = 0.0024; bottom 30%: *p* = 0.0061; **Fig. 5e**). DN2-like cells from *IL12A*-high donors trended toward higher cytotoxicity compared to *IL12A*-low donors (median = 0.136 vs 0.112, Wilcoxon rank-sum *p* = 0.084), suggesting that IL12A expression may further amplify the cytotoxic signature in DN2-like cells (**Fig. 5e**). Taken together, these validation analyses from a patient B-cell dataset demonstrate that cytotoxicity represents a genuine transcriptional program present in DN2 B cells from SLE patients, a feature that is pronounced in patients with high *IL12A* expression.

**Figure 5.**
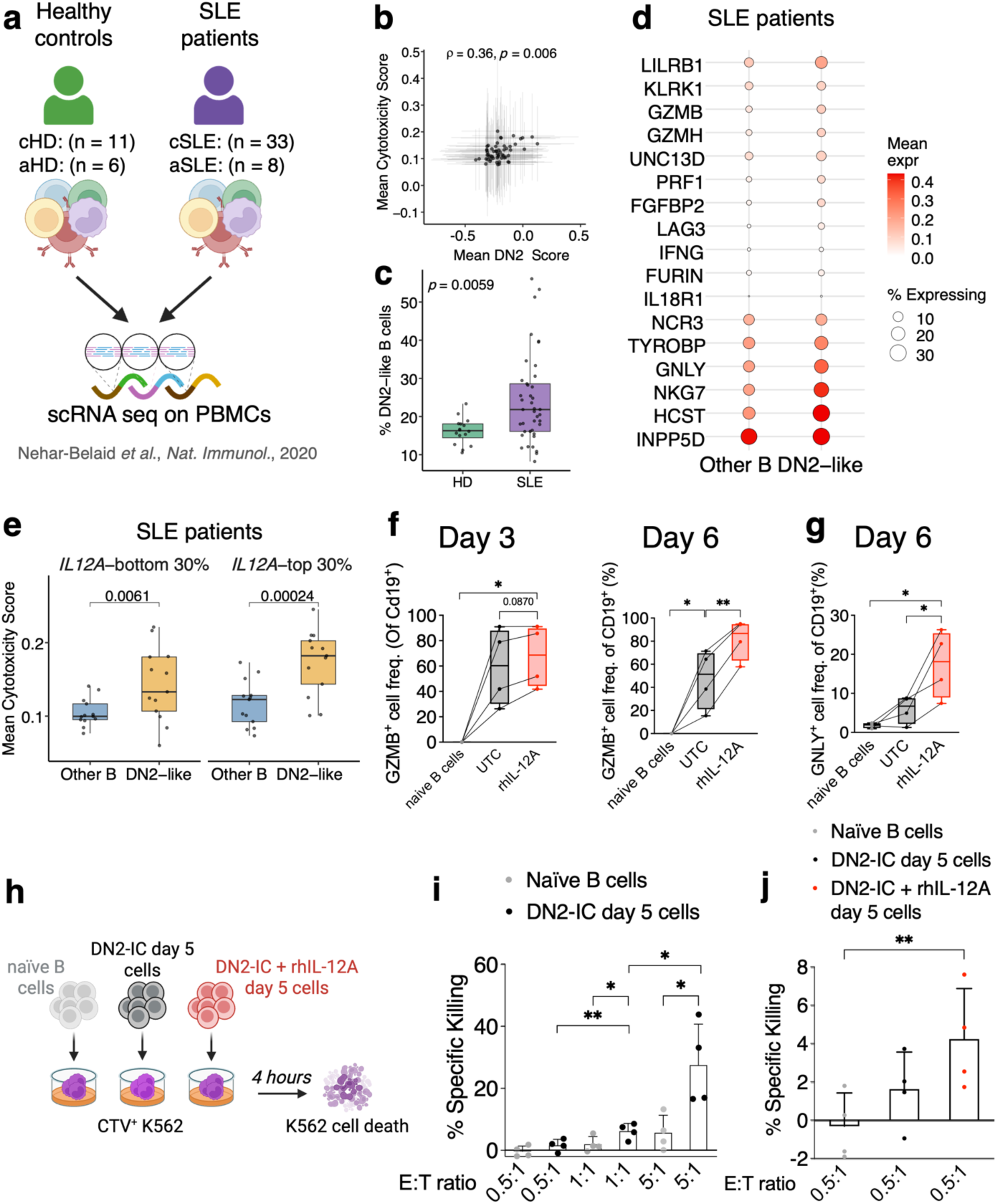
DN2 B cells inherently possess a cytotoxic program and function. **(a)** Representative scheme of the single-cell RNA-seq dataset from Nehar-Belaid et al. 2020 Single-cell RNA sequencing was performed on PBMCs isolated from children with SLE (cSLE) and matched healthy donors (cHD), as well as an independent cohort of adult SLE patients (aSLE) and adult healthy donors (aHD). **(b)** Scatter plot showing per-donor mean cytotoxicity score vs. mean DN2 score across B cells. Spearman correlation (ρ) and p-value obtained from Spearman correlation test are indicated in the figure. **(c)** Box plot comparing DN2-like cell frequencies between healthy donors (HD) and SLE patient samples. P-value from Wilcoxon rank-sum test is indicated in the figure. **(d)** Dot plot showing normalized expression of cytotoxicity-related genes in DN2-like and other B cells from the PBMCs single-cell RNA-seq data in SLE patients. **(e)** Box plot showing per-donor mean cytotoxicity score of DN2-like cells and other B cells from SLE patients. Donors are stratified by top and bottom 30th percentile of mean *IL12A* expression within SLE patients. Each point represents one donor. P-value is obtained from Wilcoxon rank-sum test. **(f)** GZMB and **(g)** GNLY expression by flow cytometry on naïve B cells stimulated with DN2-IC at the indicated days post-stimulation.**(h)** Representative scheme of the K562 killing assay: CTV-labeled K562 target cells were co-cultured for 4 hours with naïve B cells left in media with BAFF overnight (ON) as a negative control, naïve B cells activated with DN2-IC for 5 days or activated with DN2-IC and rhIL-12A. K562. K562 cell death was evaluated by FACS. **(i)** Percentage of specific killing of K562 cells co-cultured at the indicated B cell:K562 cell ratios with naïve B cells stimulated with DN2-IC for 5 days, or with naïve B cells stimulated with BAFF ON.**(j)** Percentage of specific killing of K562 cells co-cultured at a 0.5:1 ratio with naïve B cells stimulated with DN2-IC for 5 days, naïve B cells stimulated with DN2-IC and rhIL-12A, or naïve B cells stimulated with BAFF ON. Data in **(f,g)** are presented as box plots showing the median and 25th/75th percentiles; whiskers indicate the range, and individual data points represent biological replicates (donors, n = 4). Data in panels **(i-j)** are from 2 independent experimental runs which were combined and plotted together. Cell populations are calculated as a percentage of total CD19^+^ B cells. Data in **(i,j)** are presented as mean ± standard deviation. Each dot represents an individual donor *(n* = 4). Panels **(f,g)** two-tailed paired t-test; \**p* < 0.05, \*\**p* < 0.01. Panels (**i,j**) one-tailed paired t-test; \**p* < 0.05, \*\**p* < 0.01.

The expression and functional role of cytotoxic proteins in B cells remain largely unexplored, with few exceptions such as GZMB with GZMB-producing B cells exhibiting either regulatory^67,68^ or cytotoxic features^69^ in the regulation of the immune response, whereas their impact on cancer progression remains a matter of debate^69,70^. Therefore, we next sought to validate whether the transcriptional upregulation of cytotoxic genes resulted in functional protein expression. We first examined the basal cytotoxic proteomic profile of naïve B cells differentiated with the DN2-IC. We observed that DN2 B cells express IL-18R1 and LAG3 proteins by day 3 (**Extended Fig.4a**). Consistent with our transcriptomic model, this protein induction was significantly potentiated by rhIL-12A treatment by day 3 (*p* = 0.012, and *p* = 0.0002, **Extended Fig. 4a-b**) and sustained through day 6, with significant IL-12A-driven upregulation of IL-18R1 (*p* = 0.0087) and a similar trend observed for LAG3. This increase was observed in both the total B cell population (**Extended Fig. 4a-b**) and within the CD11c^+^ cells at day 6 (LAG3 *p* = 0.0037 and IL18R1 *p* = 0.0058, **Extended Fig. 4c**), while unstimulated naïve B cells remained negative for these markers. Regarding intracellular effector molecules, we observed that the DN2-inducing cocktail intrinsically drives Granzyme B (GZMB) production. This expression showed a trend toward an increase with rhIL-12A treatment at day 3 and became significant by day 6 (*p* = 0.0047), with approximately 80% of B cells expressing GZMB upon IL-12A stimulation (**Fig. 5f**). Notably, GZMB production was more pronounced and highly responsive to IL-12A within the CD11c^+^ compartment, displaying significant increases at both day 3 (*p* = 0.046), and day 6 (*p* = 0.01) compared to the non-CD11c^+^ cells (**Extended Fig. 3d-e**). Similarly, granulysin (GNLY) was upregulated by day 6 following rhIL-12A treatment (*p* = 0.045, **Fig. 5g**), with expression almost exclusively restricted to the CD11c^+^ population (**Extended Fig. 4f-g**). In contrast, perforin expression remained low and unchanged following IL-12A stimulation (**Extended Fig. 4h**), suggesting a perforin-independent, GZMB and GNLY-predominant cytotoxic profile potentiated by IL-12 signaling. To functionally validate this newly identified cytotoxic program, we assessed K562-specific killing by *in vitro*-differentiated DN2 B cells. Specifically, CTV-labeled K562 target cells were co-cultured for 4 hours with naïve B cells activated with DN2-IC for 5 days (50-80% expected DN2 B cells) or, as a negative control, with naïve B cells left in media with BAFF overnight, at various effector-to-target (E:T) ratios (0.5:1, 1:1, 5:1), or with rhIL-12A-treated *in vitro-*generated DN2 cells (**Fig. 5h**). We hypothesized that *in vitro*-generated DN2 B cells have a superior killing capacity compared to naïve B cells. Specific killing was quantified by flow cytometry using Annexin V and PI staining (**Extended Fig. 5a**). The cytotoxic effect was primarily characterized by an increase in target cells with PI-permeable membranes (Annexin V-PI+ population), which was utilized to calculate specific killing (**Fig. 5i** and **Extended Fig. 5b-c**). DN2-enriched cells exhibited killing capacity in a ratio-dependent manner, whereas naïve B cells showed absent to minimal killing (**Fig. 5i**). Furthermore, while at the lowest ratio of 0.5:1, DN2-enriched cells showed negligible killing, IL-12A treatment significantly increased killing capacity compared to the naïve condition (**Fig. 5j**). Overall, these results demonstrate that DN2 B cells possess a proteomic and functional cytotoxic profile that is potentiated by IL-12A.

### rs485499 risk allele is associated with DN2 expansion and *IL12A* upregulation in primary B cells

To functionally link our genetic findings at rs485499 within the 3q25.33 locus to the biological role of IL-12A in DN2 differentiation, we recruited healthy donors homozygous for either the risk allele (TT, *n* = 8) or non-risk allele (CC, *n* = 7). We hypothesized that B cells from risk allele carriers would exhibit increased *IL12A* expression and DN2 abundance compared with non-risk carriers. Indeed, PBMC characterization revealed that risk allele carriers exhibited higher frequency of DN2 B cells compared to non-risk carriers (*p* = 0.03, **Fig. 6a**). While the risk group showed an overall reduction in B cell compartment expansion (**Extended Fig. 6a**), frequencies of the other major B cell subsets remained comparable between the two groups (**Extended Fig. 6b-c**). We then assessed whether this genetic association effect could be recapitulated in an *in vitro* differentiation assay under DN2-skewing conditions. Naïve B cells from the same donors were cultured with the DN2-IC, and DN2 B cell frequency was analyzed at days 1, 3, and 6. Risk allele carriers exhibited a significant increase in DN2 B cell generation at days 1 (*p* = 0.007) and 3 (*p* = 0.046), with a similar trend at day 6 (**Fig. 6b**). No significant changes were detected in the overall DN population (**Extended Fig. 6d**). By day 6, we also observed a significant increase in plasmablast (PB) differentiation in risk-allele carriers (*p* = 0.005, **Fig. 6c**). Given that DN2 B cells represent a pre-antibody secreting cell state^14,15^, we believe that the enhanced plasmablast differentiation at day 6 may reflect the elevated DN2 generation in risk allele carriers at earlier time points. Indeed, we observe a positive correlation between DN2 abundance at day 1 (*p* = 0.09, *r* = 0.46) and day 3 (*p* = 0.0063, *r* = 0.47) and plasmablast abundance at day 6 (**Extended fig. 6e**).

**Figure 6.**
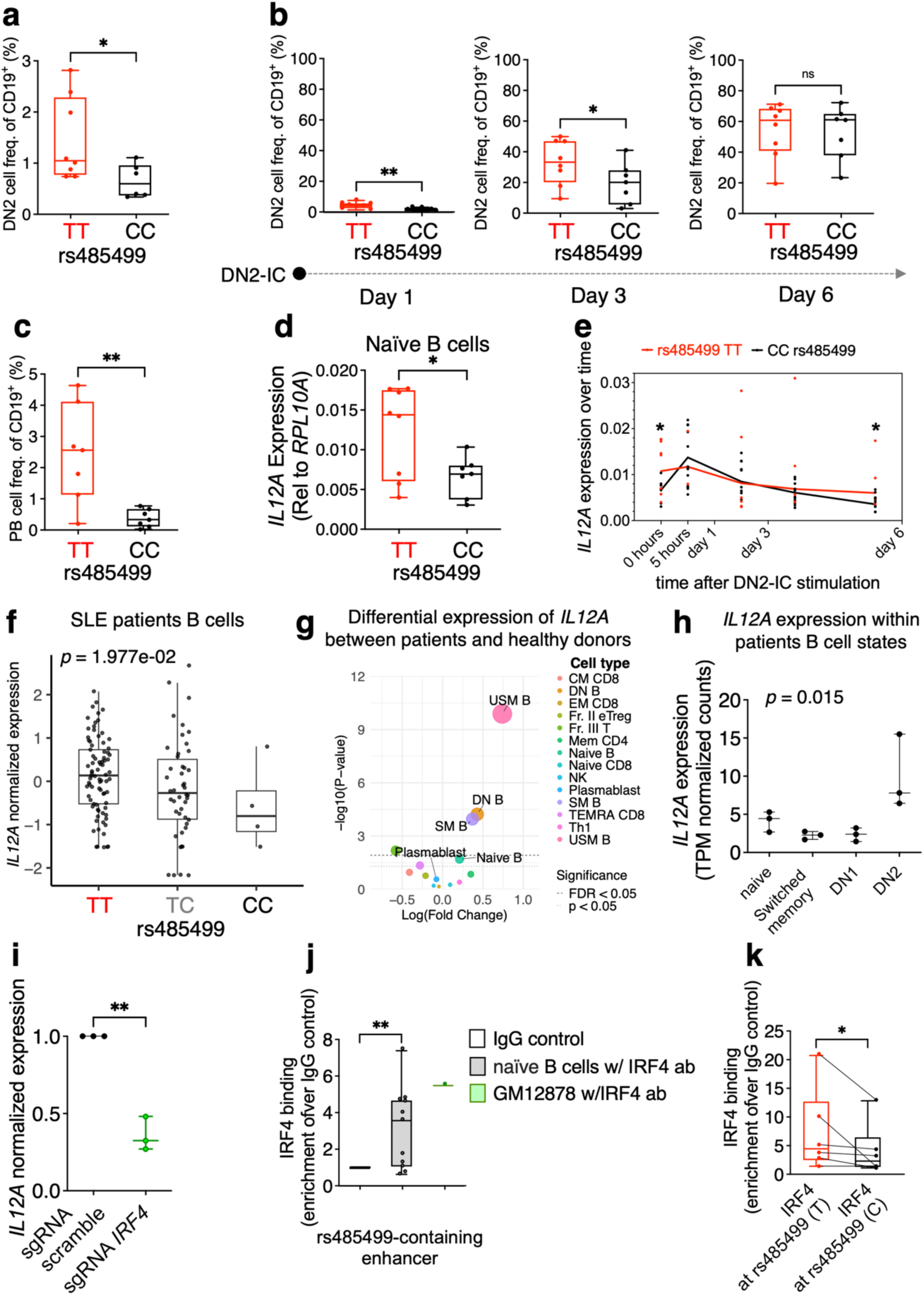
rs485499 risk allele is associated with DN2 expansion in PBMCs, increased *IL12A* expression and enhanced IRF4 binding in naïve B cells. **(a)** DN2 B cell percentage in peripheral blood compared between donors homozygous for the rs485499 risk allele (TT, *n* = 8) and non-risk allele (CC, *n* = 7). **(b)** DN2 B cell and **(c)** plasmablast percentages in naïve B cells from risk and non-risk allele carriers stimulated in vitro with DN2-IC at the indicated time points. (d) *IL12A* gene expression in naïve B cells from risk versus non-risk allele carriers. (e) *IL12A* expression kinetics in naïve B cells from risk allele carriers (in red) and non-risk carriers (black) stimulated with DN2-IC. **(f)** eQTL analysis showing the association between rs485499 and *IL12A* expression in B cells from SLE patients (*n* = 70). **(g)** Bubble plot illustrating the differential expression of *IL12A* between healthy donor cells and inactive SLE patient cells derived from Supplementary Table S2 of Nakano, Masahiro et al. Cell 2022. The x-axis represents the log (fold change) in expression, while the y-axis shows statistical significance as -log10(P-value). The horizontal dashed line indicates the threshold for FDR < 0.05, and the dotted line represents *p* < 0.05. Each bubble is color-coded by cell type. The size of each bubble corresponds to the magnitude of the fold change and statistical weight for each subset. In panel h each dot represents a donor. **(h)** Box plot showing Transcripts Per Million (TPM) for *IL12A* across indicated B cell subsets in SLE patients. Data were obtained from the published dataset by Jenks et al. Immunity 2018. (i) *IL12A* expression at day 6 following *IRF4* CRISPR KO in naïve B cells stimulated with DN2-IC. **(j)** IRF4 CUT&RUN followed by qPCR showing IRF4 enrichment at the rs485499-containing region in naïve B cells, using GM12878 cells as a control. **(k)** Allele-specific binding of IRF4 at the rs485499 T and C alleles using TaqMan assay with allele specific probes. In panels **(a-d and f)** and **(i-k)**, data are presented as box plots showing the median and 25th/75th percentiles; whiskers indicate the range, and individual data points represent biological replicates (3 to 11 donors per group). Data in panels **(j-k)** represent combined results from 3–4 independent experimental runs, and data in panel **i** represent combined results from 2 independent experimental runs. In panel **f** each dot represents a donor-visit SLE sample, x-axis and y-axis represent genotype and mean normalized expression of *IL12A* in B cells, respectively. Cell populations in **(a-c)** are calculated as a percentage of total CD19^+^ B cells. Gene expression is normalized to *RPL10A* (**d**) and expressed relative to time 0 of stimulation (naïve B cells) in **(e)** or to the scramble negative control **(i)**. In **(j, k)**, IRF4 values are normalized to IgG control values using the ΔΔCt method. Panels **(a-d)**: One-tailed Mann-Whitney U test \**p* < 0.05, \*\**p* < 0.01. Panels **(e, i, j)**: Two-tailed paired t test, \**p* < 0.05, \*\**p* < 0.01. Panel **(k)**: One-tailed paired t-test. \**p* < 0.05. Panel **(f)** p-value was obtained from linear-mixed model with donor as random effect testing eQTL in B cells, adjusted for ancestry with genetic principal components, technical effects with transcriptional principal components and sex. Panel **(h)**: each dot represents a patient sample. Kruskal-Wallis test. Differences were considered statistically significant at *p* ≤ 0.05.

To confirm that DN2 expansion in risk carriers was linked to increased *IL12A* expression, we measured *IL12A* gene expression by qPCR in naïve B cells from the same individuals under identical experimental conditions. In line with our hypothesis, in untouched naïve B cells, we observed higher expression levels of *IL12A* in risk allele carriers compared to non-risk carriers (*p* = 0.046, **Fig. 6d**). Moreover, this increased *IL12A* expression was also evident following *in vitro* differentiation under DN2-inducing conditions, with significant differences emerging by day 6 (*p* = 0.021, **Extended Fig. 6f**). Supporting the role of IL12A in DN2 expansion, we observed a trend toward a positive correlation between *IL12A* expression in naïve B cells and DN2 B cell abundance in PBMCs (*r* = 0.19, *p* = 0.057) and a stronger significant positive correlation (*r* = 0.59, *p* = 0.0008) between *IL12A* expression in naïve B cells and DN2 expansion *in vitro* at day 1 (**Extended Fig. 6g-h**). Furthermore, when examining *IL12A* expression kinetics over time, we observed that in non-risk carriers, *IL12A* expression peaked early and declined thereafter (**Fig. 6e and Extended Fig. 6i**) consistent with our previous observations (**Fig. 3a**) and prior reports^13^. In contrast, risk allele carriers exhibited a sustained *IL12A* expression pattern over time (**Fig. 6e and Extended Fig. 6i**), suggesting prolonged *IL12A* production during DN2 B cell differentiation.

To determine if the rs485499 risk allele is similarly associated with increased *IL12A* expression in SLE patients B cells, we leveraged an unpublished PBMC scRNA-seq dataset of 70 genotyped SLE patients (ongoing project that will be published elsewhere). eQTL analysis in B cells confirmed that the risk allele is significantly associated with increased *IL12A* expression within this patient cohort (*p* = 1.97e-02, **Fig. 6f**). Moreover, bulk RNA-seq data on peripheral immune cells from a published study^71^ revealed that multiple B cell subsets from SLE patients — including the double negative (DN) population — exhibited a significant upregulation of *IL12A* compared to healthy controls cells, which was not the case for other immune cell types (**Fig. 6g**). Analysis of sorted peripheral blood B cell subsets from a publicly available dataset^14^ revealed significant differential expression of *IL12A* across distinct B cell compartments (Kruskal-Wallis, *p* = 0.043). Notably, *IL12A* expression was upregulated in DN2 B cells compared to DN1, resting naïve, and switched memory populations, pointing to DN2 cells as a primary source of B cell-derived *IL12A* in SLE (**Fig. 6h**). To further validate this B-cell intrinsic source of *IL12A* at single-cell resolution, we interrogated the Accelerating Medicines Partnership SLE Network peripheral blood atlas^72^. Single-cell transcriptomic analysis showed that DN2/ABCs (cluster bl-B2. *FCRL5*+ *ITGAX*+ ABC) exhibit the highest enrichment of *IL12A* transcripts among the B cell subsets (padj< 4.67e-62), followed by the naive B-cell compartment (bl-B0. *CXCR5*^high^ Naive, padj< 5.58e-14)^72^. Finally, validation using the summary statistics of the UK Biobank atlas of the plasma proteome in >50,000 individuals^73^ revealed a highly significant positive association between plasma IL-12 levels and SLE (*p* < 5.79e-40). This evidence aligns with previous clinical reports^74,75^ and reinforces the relevance of the IL-12 pathway in lupus pathogenesis.

Overall, these results suggest that rs485499 risk allele drives DN2 B cell expansion and increased *IL12A* expression in healthy donors and SLE patients, and highlights B cells including DN2s as critical *IL12A* producers in SLE.

### rs485499 risk allele increases transcription factor IRF4 binding

Finally, to investigate the mechanism underlying rs485499-mediated allele-specific regulation of *IL12A*, we first examined transcription factors predicted to bind the enhancer region harboring rs485499. ChIP-seq data performed in the GM12878 B cell line^76^ and in activated memory primary B cells^77^ shows this locus is occupied by multiple transcription factors including IRF4 (**Extended Fig. 7a**). Furthermore, rs485499 resides within a predicted IRF4 binding motif (**Extended fig. 7b, Extended Table 3 and Extended Methods**)^78^. IRF4 is a master regulator of plasma cells differentiation during extrafollicular response^26,29,77^, and DN2 cells display elevated IRF4 transcript levels along with enrichment of predicted IRF4 target gene expression^14^. Further, IRF4 protein expression was upregulated during differentiation of naïve B cells with the DN2-IC (**Extended fig. 7c**). Resting naïve B cells express low, yet detectable, levels of IRF4 protein (**Extended fig. 7d**). Moreover, *IRF4* knockout in GM12878 cells reduces *IL12A* expression^79^. Together, these findings point to IRF4 as a candidate transcription factor regulating *IL12A* during DN2 B cell differentiation. We first sought to confirm the functional role of IRF4 on *IL12A* regulation in primary cells. We therefore performed CRISPR–Cas9–mediated knockout of *IRF4* in naïve B cells stimulated with the DN2-IC. Given that *IRF4* gene expression peaks approximately 4 hours after stimulation, we introduced CRISPR editing 1-hour post-stimulation to maximize knockout efficiency, achieving ∼40% editing efficiency (**Extended Fig. 7e**). *IRF4* knockout resulted in reduced *IL12A* expression in primary B cells compared to the scramble control at 6 days post-activation with DN2-IC (*p* = 0.0095, **Fig. 6i**), consistent with findings in GM12878 cells^79^.

To directly assess IRF4 binding, we performed Cleavage Under Targets and Release Using Nuclease (CUT&RUN) followed by qPCR in naïve B cells. We confirmed antibody specificity and workflow integrity via IRF4 enrichment and H3K4me3 occupancy at a known MYC super-enhancer^80^ (**Extended Fig. 7f**). We observed significant IRF4 enrichment at the rs485499-containing enhancer compared to the IgG control (**Fig. 6j**), a finding further validated using GM12878 cells as a positive control (**Fig. 6i and Extended Fig. 7a**). IRF4 rs485499 allele-specific binding was evaluated using a TaqMan genotyping assay in donors heterozygous for rs485499. Based on the association of the risk allele to increased *IL12A* production and our data supporting IRF4 dependent regulation of *IL12A* expression, we hypothesized that IRF4 would preferentially bind the risk allele (T) over the non-risk allele (C). Consistent with this hypothesis, among donors with positive IRF4 enrichment we observed significantly higher IRF4 enrichment on the T allele (*p*= 0.048 **Fig. 6k**). Together, these results support a model in which IRF4 preferentially binds to the rs485499 risk allele, driving upregulation of *IL12A* expression in naïve B cells during differentiation into DN2 B cells.

## DISCUSSION

SLE pathogenesis is driven by complex genetics, but linking non-coding risk variants to specific pathogenic cellular phenotypes remains a major bottleneck. In this study, we bridge this gap by demonstrating that a common SLE genetic risk variant at the 3q25.33 locus directly drives the expansion of a pathogenic B cell subset. By integrating *in silico* predictions, CRISPR-based validations, and primary human cell assays, we identify rs485499 as a likely causal variant and *IL12A* as its target gene. We demonstrate that the risk allele upregulates *IL12A* and is associated with expanded DN2 B cells in periphery. Meanwhile we show that IL-12A not only promotes the expansion of DN2 B cells but arms them with a previously unrecognized cytotoxic program. Together, these findings establish a direct variant-to-function link that defines a new pathogenic axis in SLE.

Resolving the functional consequences of non-coding GWAS variants requires navigating cell-state-specific regulatory dynamics^81,82^. Using CRISPRi and adenine base editing, we establish the rs485499 region as a causal enhancer controlling *IL12A* expression. Crucially, the regulatory impact of this variant is highly context-dependent, as it has also been shown for other autoimmune loci^83,84^. While previous eQTL data from LCLs suggested the risk allele decreases *IL12A* expression^42,43^, and our base editing in LCLs confirmed this, our functional validation in primary B cells indicate the opposite: the risk allele is associated with increased *IL12A* expression during early DN2 differentiation in healthy donors, as well as in B cells from SLE patients. Mechanistically, this increase is driven by the transcription factor IRF4, which preferentially binds the rs485499 risk allele in naïve B cells to promote *IL12A* upregulation. This observation highlights the necessity of validating non-coding variants within the specific immune cell states relevant to disease pathogenesis.

While B cells are a known primary source of *IL12A* transcripts among immune cell types ^53,54^, the precise role of B cell-derived IL-12 has remained poorly defined. Recent work identified a B cell-intrinsic IL-12/IFN-γ feed-forward loop driving plasmablast differentiation^34^; our data expand this paradigm by placing IL-12 as a central driver of the DN2 subset. In our *in vitro* model, *IL12A* is transiently upregulated early after activation, and its CRISPR-mediated disruption significantly impairs DN2 expansion. Conversely, providing exogenous IL-12A increases both IFN-γ production and DN2 differentiation— an effect explicitly dependent on IL-12, rather than IL-35, signaling. Because rs485499 risk-allele carriers exhibit a sustained, rather than transient, *IL12A* expression profile over B cell activation, this prolonged production likely potentiates the IL-12/IFN-γ circuit. Supported by our observation that multiple B cell states in SLE, including the DN compartment, exhibit elevated *IL12A* with DN2 B cells showing the highest levels, we propose that this continuous signaling contributes to the pathogenic expansion of the DN2 population.

Beyond their established role as pre-plasma cells^15,85,86^, our transcriptomic and functional analyses reveal that DN2 B cells possess intrinsic cytotoxic capabilities. The expression of cytotoxic molecules in B cells has been sporadically reported, such as GZMB in regulatory contexts or under incomplete T cell help^67–69^ and GNLY in antibacterial responses^87^. However, their concerted pathogenic role in autoimmunity remains largely undefined. In both our *in vitro* models and SLE patient cohorts, DN2 cells exhibit a distinct cytotoxic transcriptional signature, which we show is responsive to IL-12 signaling. Based on our protein-level profiling, we propose that this DN2-mediated cytotoxicity is perforin-independent—as has been shown for other GZMB-expressing B cells^69^—relying instead on a synergistic GZMB/GNLY program to induce rapid cell membrane permeabilization, consistent with the established cooperative actions of these lytic molecules^88^. Because DN2 cells are known to accumulate in lupus-affected kidneys and correlate with renal damage^86,89,90^, this newly identified cytotoxic program provides a new possible mechanism: rather than acting solely as autoantibody precursors, these cells may have the capability of directly mediating localized tissue destruction.

While these findings provide a comprehensive model of the IL-12/DN2 axis, several limitations warrant consideration. First, due to the current technical constraints of achieving highly efficient base editing in primary resting B cells, our rs485499 editing was performed in a B cell line, and it resulted in a bystander edit at an adjacent adenine. However, because this adjacent position is not polymorphic in the human population, and our primary B cell data demonstrate allele-specific IRF4 binding, the combined evidence strongly positions rs485499 as the causal variant. Still, we cannot entirely rule out synergistic contributions from other variants in high linkage disequilibrium at the 3q25.33 locus. Finally, while the K562 tumor cell line is a standard *in vitro* model for assessing NK cell cytotoxicity due to its lack of MHC class I expression, future studies are needed to elucidate how B cells recognize these cells. Prior research indicates that B cells can mediate cancer cell killing through the engagement of multiple TNF superfamily ligands^91^, many of which—including TNF, TRAIL and OX40L—are expressed by DN2 B cells^13,92,93^ which concurrently lack multiple negative regulators of the TNF family^14^. Given that K562 cells natively express a broad repertoire of TNF-family receptors^94,95^, this signaling node represents a plausible mechanism for DN2 B cell-mediated target engagement. These receptor-ligand interactions could serve as a hypothetical mechanism to trigger or enhance the perforin-independent GZMB/GNLY delivery program. Furthermore, utilizing autologous tissue models will be required to definitively establish the mechanism of receptor-ligand interactions and subsequent DN2-mediated damage in SLE target organs.

Ultimately, defining this variant-to-function mechanism provides a compelling rationale for precision therapeutic interventions in SLE. Because drug targets supported by human genetic evidence are historically more likely to succeed clinically^96,97^, our findings offer a framework to re-evaluate previous therapeutic setbacks. Specifically, the failure of ustekinumab (anti-IL-12/23) in phase III SLE trials^98^ may reflect a lack of subset-specific patient stratification rather than a flawed biological target. While individual non-coding GWAS variants confer only modest disease risk, rs485499 serves to unmask a broader pathogenic axis that culminates in DN2 B cell expansion. Notably, individuals of African ancestry exhibit inherently increased DN2 frequencies^14,99,100^but comprised only 8% of the ustekinumab trial participants^98^. Therefore, stratifying future clinical cohorts based on the magnitude of DN2 expansion could rescue the clinical utility of IL-12 inhibition. Furthermore, by providing genetically linked evidence that defines DN2 B cells as pathogenic drivers of SLE, this work strongly supports emerging precision cellular therapies. As the field advances toward highly selective immune depletion, targeting the DN2 subset with engineered CAR-T cells or bispecific antibodies^101–103^ offers a prime opportunity to eliminate disease-driving cells while preserving baseline humoral immunity.

## MATERIALS AND METHODS

### Human cells collection

Peripheral blood collar samples were collected from anonymous donors at the Boston Children’s Hospital Blood Donor Center under IRB-P00046424. Peripheral blood mononuclear cells (PBMCs) were isolated by Ficoll-Paque density gradient centrifugation.

### rs485499 genotyping

For rs485499 genotyping, Genomic DNA was extracted from either 100 μL of PBMCs or a minimum of 50,000 GM12878 cells using the GeneArt™ Genomic Cleavage Detection Kit (#A24372, Thermo Fisher Scientific), following the manufacturer’s instructions. The genomic region encompassing rs485499 was PCR-amplified using specific primers (Forward1: CCAGACCCTGTTTTACTACTTCA, Reverse1: TGAGGAGAGGCAGTTAAACCA, Forward2: CAGTGGCCCATACCTGTAATC, Reverse2: CTCTGTCTCCCAAAGTGCTAAA). Unpurified PCR products or genomic DNA were submitted to Azenta Life Sciences or Quintarabio for Sanger sequencing. Sequencing chromatograms were analyzed using SnapGene, and genotyping/editing efficiency was quantified using the EditR tool. Among the 15 donors used in Figure 4, 8 were homozygous for the T allele (TT), and 7 were homozygous for the C allele (CC) at rs485499.

### Naïve B cell purification and stimulation with the DN2-inducing cocktail

Naïve B cells were isolated from peripheral blood mononuclear cells (PBMCs) using the Naïve B Cell Isolation Kit (Stemcell Technologies). Cell purity (>90%) was confirmed via flow cytometry (FACS). To induce differentiation into the DN2 B cell state, naïve B cells were plated at a density of 1 million cells/mL in complete RPMI medium and stimulated with a DN2-inducing cocktail consisting of 10 ng/mL BAFF (Biolegeng, #559602), 10 ng/mL IL-21 (Biolegeng, #571202), 2.12 ng/mL IL-2 (#589102, Biolegend) 20 ng/mL IFN-γ (#570204, Biolegend) 1 μg/mL R848 (Invivogen, #TLRL-R848), 5 μg/mL anti-IgG/IgM (Jackson ImmunoResearch, # 109-006-127) and 5 μg/mL anti-IgA (Jackson ImmunoResearch, #109-005-011). Cells were cultured for 6 days and were restimulated on day 3. When specified recombinant human IL-12A (#11209-IL, R&D Systems) was added to the DN2-inducing cocktail at the concentration of 200ng/ml, while ustekinumab biosimilar (#SIM0020, Bioxcell) was added at 10ug/ml and kept for all the stimulation protocol.

### RNA extraction and quantitative PCR (qPCR)

Total RNA was extracted using the miRNeasy Micro Kit (#217084, Qiagen), and cDNA was synthesized with the AffinityScript cDNA Synthesis Kit (#200436, Agilent). Quantitative PCR (qPCR) was performed using Brilliant II SYBR® Green QPCR Master Mix (#600828, Agilent) on a QuantStudio 3 Real-Time PCR System (Applied Biosystems). Primer sequences for all targets are provided in **Extended Table 4**. Gene expression was normalized to *GAPDH* or *RPL10A* and expressed relative to the control sample using the ΔΔCt method.

### Flow cytometry

For surface protein staining, cells were washed with PBS and incubated with 100 μL of Zombie Aqua™ viability dye (#423101, Biolegend) for 10 minutes at room temperature (RT), according to the manufacturer’s instructions. Following an additional wash with PBS containing 2% FBS (FACS buffer), cells were stained with 100 μL of an antibody cocktail for 30 minutes at RT. For intracellular protein staining (GZMB, PRF1, and GNLY), cells were incubated with Brefeldin A (Brefeldin A Solution, #00-4506-51, eBioscience™) at 1X for 4 hours at 37 °C. After incubation, cells were washed with PBS and stained using the Foxp3 / Transcription Factor Staining Buffer Set (#00-5523-00, eBioscience™) following the manufacturer’s instructions. Antibodies conjugated against the proteins of interest, as well as appropriate isotype controls, were used. Samples were acquired on a BD LSRFortessa™ flow cytometer (BD Biosciences). Instrument performance was verified using single-stained compensation beads for experiments performed at a single time point or Rainbow Calibration Particles (Spherotech) when data were collected across multiple time points. Data were collected using FACSDiva software (BD Biosciences) and analyzed with FlowJo software version 10.5.3 (BD Biosciences). Complete antibody panels and the gating strategy used to analyze Flow cytometry data are illustrated in **Extended tables 5-6.**

### Colocalization analysis

SLE GWAS summary statistics were downloaded from the GWAS Catalog (GCST007400)^7^, and lifted over from the GRCh37 to the GRCh38 reference assembly using the *+liftover* plugin in bcftools v1.22^104^. A locus surrounding the *IL12A* gene was defined using a ±500 kb window centered on the lead GWAS variant. Summary statistics for gene expression quantitative trait loci (eQTLs) were acquired from the eQTL Catalogue (accessed in March 2026)^41,105^. For all 280 available studies, data within the *IL12A* genomic window were extracted using the tabix method. Bayesian colocalization assuming a single causal variant per locus was performed using the coloc R package v5.2.3 to assess the probability of a shared causal variant between the SLE GWAS and *IL12A* eQTL signals using default prior probabilities^106^.

### In silico prediction of putative enhancer-associated SNPs located within active or poised regulatory elements

Risk variants within the SLE 3q25.33 locus were obtained from the Langefeld et al. genome-wide association study (GWAS)^7^ and cross-referenced with the 1000 Genomes Project database. Variants meeting genome-wide significance (*p* < 1e-5) and exhibiting high linkage disequilibrium (r2 > 0.8) with the lead causal variant were prioritized (*n* = 63). To evaluate the regulatory potential of these candidates, a 1 Mb window was established around each prioritized variant. Epigenomic and functional genomic datasets for the GM12878 lymphoblastoid cell line were integrated to identify active transcriptional regulatory elements (TREs). Specifically, active TREs were defined using global run-on sequencing (GRO-seq) data^48^ processed via the dREG gateway. This dataset was complemented by ENCODE functional genomic annotations^47^, including chromatin immunoprecipitation sequencing (ChIP-seq) profiles for key histone modifications (H3K4me1: ENCFF921LKB, H3K4me3: ENCFF295GNH, and H3K27ac: ENCFF816AHV); ATAC-seq accessibility profiles (ENCFF470YYO); and DNase I hypersensitivity sites (DNase seq, ENCSR000EMT). Variants overlapping any of these functional regulatory features were assigned a binary score of 1. To specifically enrich for distal regulatory elements, variants overlapping ChromHMM-defined promoter regions were systematically excluded in the initial filtering tier. This filtering strategy narrowed the candidate pool to 36 variants categorized as “enhancer” variants (**Fig.1b** and **Extended Table 2**). Through this approach, rs485499 was identified as a top candidate causal variant overlapping the greatest number of chromatin modifications predictive of regulatory regions such as enhancers and these were: H3K4me1, H3K27ac, ATAC-seq DNase-seq and GRO-seq.

### CRISPR-Cas9 Knockout in Primary B Cells

Naïve B cells (1–1.5 × 10⁶) were stimulated with the DN2-inducing cocktail for 1 hour prior to nucleofection. After stimulation, cells were collected, washed with PBS, and resuspended in P3 Primary Cell Nucleofector™ Solution (#V4XP-3032, Lonza) following the manufacturer’s instructions. For RNP complex formation, a 4:1 molar ratio of synthetic single-guide RNA (sgRNA) to Cas9 protein (60005575-2, EditCo) was used. The sgRNAs targeting *IL12A, EBI3*, and *IRF4* were purchased from Synthego as a chemically modified, three-guide multiplex Gene Knockout Kit. A non-targeting scrambled sequence (scramble) was used as a control. The sgRNA sequences are provided in **Extended Table 7.** The RNP complexes were incubated at room temperature for 15 minutes before use. Nucleofection was performed using the EH115 program on the Lonza 4D-Nucleofector™ system. Immediately following nucleofection, 100 µL of pre-warmed complete RPMI medium supplemented with 10% FBS, penicillin/streptomycin, and DN2-inducing cocktail was added to each cuvette and incubated for 15 minutes at room temperature. Cells were then transferred to individual wells of a 48-well plate containing 300 µL the same media. After 3 days, cells were used for downstream analyses.

### CRISPR technologies in GM12878

GM12878 lymphoblastoid B cells (LCLs)—a human Epstein-Barr virus (EBV)-transformed ENCODE Tier 1 cell line—were obtained from the Coriell Institute. Cells were maintained in RPMI supplemented with 10% fetal bovine serum, 1% penicillin-streptomycin, and 1% L-glutamine, at a density of 2 × 10⁵ cells/mL at 37°C in a humidified incubator with 5% CO₂. GM12878 cells stably expressing the Cas9 CRISPR system were kindly provided by the laboratory of Benjamin Gewurz. To establish stable expression of the CRISPR interference (CRISPRi) system in GM12878 cells, lentiviral particles were generated using either the lentiCRISPRi(v1)-Blast plasmid (Addgene #170067, expressing dCas9-KRAB) or lentiCRISPRi(v2)-Blast, following established protocols^51^. sgRNA (**Extended table 7**) targeting the regulatory region encompassing rs485499 were designed using CRISPRpick (Broad Institute), selecting guides with high predicted on-target efficiency and minimal off-target potential. The Scramble sgRNA was used as a control. Nucleofection was performed using the SE Cell Line 4D-Nucleofector® X Kit (#V4XC-1032 Lonza) according to the manufacturer’s instructions. Briefly, 300,000– 500,000 GM12878 cells were resuspended in SE solution and nucleofected with 6 µL of 30 µM sgRNA using program DN-100. Immediately post-nucleofection, cells were transferred to pre-warmed RPMI medium and cultured for 72 hours to enable effective gene silencing. qPCR was performed as described above. *IL12A* transcript levels were normalized to *GAPDH* and expressed relative to the scramble control using the ΔΔCt method.

### Adenine base editor (ABE) technology

GM12878 were maintained in RPMI supplemented with 10% fetal bovine serum, 1% penicillin-streptomycin, and 1% L-glutamine, at a density of 2 × 10⁵ cells/mL at 37°C in a humidified incubator with 5% CO₂. To establish stable expression of the adenine base editor ABE8e system in GM12878 cells, lentiviral particles were generated using the pRDA_876 plasmid (a gift from the Broad Institute), following established protocols^51^. A single-guide RNA (sgRNA) targeting the deamination of rs485499 (A base) was manually designed based on previously published protocols^107^ (**Extended Table 7**). The Scramble sgRNA was used as a control. Nucleofection was carried out as described above. Immediately after nucleofection, cells were transferred to pre-warmed complete RPMI medium and cultured for 72 hours to allow for gene editing activity. Rs485499 genotyping and *IL12A* qPCR were performed as described above.

### Single-cell RNA-seq library preparation

Naive B cells from five healthy donors were stimulated with a previously described DN2-inducing cocktail for 1, 3, and 6 days. To support sustained activation and cell viability, cultures were restimulated on day 3. For each donor, cells were split into two conditions: DN2 stimulation alone (unstimulated treatment control, UTC) or DN2 stimulation supplemented with recombinant human IL-12A (rhIL-12A, 200 ng/mL). At each time point, cells were harvested and stained with Hash Tag Oligonucleotides (HTOs) using human TOTAL-Seq C Repertoire (5’) (**Extended Table 8**) following manufacture instructions, to mark cells from each donor and condition. For each time point, all 10 samples were pooled, and live CD19⁺ cells were sorted before cell RNA sequencing. Each time point was processed as an independent library. Droplet encapsulation was performed in-house using the 10x Chromium platform, and cDNA conversion following the manufacturer’s Chromium GEM-X Single Cell 5’ Reagent Kits v3 protocol. Library construction was performed simultaneously once all samples were collected to minimize batch effects. Paired-end 5’ sequencing was performed by Azenta Life Sciences using an Illumina platform (2 x 150bp, 950 ∼ Gb output).

### Single-cell RNA-seq data processing

Raw sequencing data were processed using CellRanger (v4.0.12), and filtered feature-barcode matrices were loaded in Python (v3.10.18) using Scanpy (v1.11.0). Gene expression (GEX) and hashtag oligo (HTO) modalities were separated per library. Doublet detection was further performed with Scrublet (v0.2.3). Sample demultiplexing was carried out in R (v4.4.3) using HTODemux in Seurat (v5.3.0), and cells classified as doublets by either HTO demultiplexing or high Scrublet scores were excluded. Cells were further filtered to retain those with 500-7,500 detected genes and mitochondrial gene content below 15%. Cell type annotation was performed using CellTypist (v1.7.0)^64^ with the Immune_All_Low model and majority voting; non-B cells identified by majority_voting_low were removed as likely contamination.

For dimensionality reduction and further QC, counts were first normalized. Mitochondrial, ribosomal, immunoglobulin, T cell receptor, HLA loci and sex-linked genes, were excluded prior to variable feature selection. Highly variable genes were identified per library and their union was used for scaling and PCA. UMAP embedding was computed using the top 20 principal components with cosine distance metric and 30 nearest neighbors with umap R package (v0.2.10.0). Shared nearest neighbor graphs were constructed and Leiden clustering was performed. Cluster quality was assessed by inspecting distributions of QC metrics, HTO classification, and Scrublet scores across clusters. Two iterative rounds of cluster-based filtering were applied, removing clusters enriched for low gene counts, HTO-negative or doublet-classified cells. Cluster marker genes were identified manually and using genesorteR (v0.4.3) to guide final cell type assignment. After all filtering steps, 50,883 cells were retained for downstream analysis.

### Pseudobulk differential expression and pathway analysis

For differential expression analysis, single-cell counts were pseudobulked by summing raw counts per donor per condition per timepoint, yielding 30 pseudobulk samples (5 donors × 2 conditions × 3 timepoints). Genes were retained if they had at least 10 counts in at least 10 of the 30 samples. Pseudobulk counts were normalized to log2(CPM+1). Differential expression between rhIL-12A-stimulated and untreated conditions was tested separately at each timepoint using linear mixed-effects models (LMM) implemented in lme4 (v1.1-37) and lmerTest (v3.1-3). For each gene and timepoint, a full model including stimulation condition and log-transformed library size as fixed effects and donor as a random effect was compared against a null model without the condition term via likelihood ratio test. Within each timepoint, genes were independently filtered to require at least 10 counts in at least 4 of the 10 samples before testing. P-values were corrected for multiple testing using the Benjamini-Hochberg method, and genes with FDR < 0.05 were considered differentially expressed.

Gene set enrichment analysis was performed using fgsea (v1.32.4) with genes ranked by LMM t-statistic. Gene sets were drawn from the MSigDB Hallmark collection and GO Biological Process collection retrieved via msigdbr (v25.1.1), comprising 7,633 pathways in total. Pathways with fewer than 15 or more than 500 member genes were excluded. Statistical significance was assessed using the fgseaMultilevel algorithm, and pathways with FDR-adjusted p-value < 0.05 were considered significant. Normalized enrichment scores (NES) for immune and cytotoxicity-relevant pathways were visualized as a heatmap across timepoints using pheatmap (v1.0.12), with gene-level t-statistics for pathway member genes displayed for pathways of interest.

### Patient validation dataset processing

To validate findings in patient-derived data, we reanalyzed a published scRNA-seq dataset of peripheral blood mononuclear cells (PBMCs) from SLE patients and healthy controls^21^. Raw sequencing data were reprocessed through CellRanger, and filtered feature-barcode matrices from 57 samples were loaded in Python (v3.10.18) using Scanpy (v1.11.0). Doublet detection was performed per sample using Scrublet (v0.2.3). The resulting 385,641 immune-enriched cells were imported into R (v4.4.3) using Seurat (v5.3.0) for quality control. Cells were retained based on the following thresholds: between 500 and 5,000 detected genes, mitochondrial gene content ≤10%, ribosomal gene content ≥10%, and hemoglobin gene content ≤1%. After filtering, 356,074 cells were retained. Counts were normalized, and highly variable genes were identified per sample (top 500 per sample, union = 1,515 genes) after excluding mitochondrial and ribosomal genes. Data were scaled and reduced to 20 principal components. Harmony (v1.2.3) batch correction was applied across samples. UMAP embedding, SNN graph construction, and Leiden clustering were performed on the Harmony-corrected embeddings. Clusters enriched for canonical B cell markers were selected, resulting in 36,538 B cells for downstream analysis.

### DN2 score and cytotoxicity score definition in B cells

To assess DN2 identity and cytotoxic potential in B cells, we calculated gene expression scores for each cell using log-normalized counts. A DN2 score was computed as the mean expression of positive DN2 markers (*FCRL5, ZEB2, DUSP4, TBX21, ITGAX, SOX5,PDCD1, ZAP70, TNFRSF1B, FGFR1*) minus the mean expression of negative markers (*CR2, IL6, FCER2, CXCR5*), where the markers were derived from a curated set of genes identified through differential expression analysis of published sorted DN2 B cells^19^ combined with established DN2-associated genes from the literature^14^. Cells with a DN2 score above 0.05 were classified as DN2-like and the remainder as non-DN2. A cytotoxicity score was computed as the mean expression of a curated gene set derived from 1) the leading-edge genes of the significantly enriched cytotoxicity-related pathways in our pseudobulk GSEA analysis (leukocyte-mediated cytotoxicity (GO:0001906), natural killer cell activation (GO:0030101), natural killer cell mediated cytotoxicity (GO:0002228), regulation of immune effector process (GO:0002697), 2) the differentially expressed genes from day 6 with cytotoxicity functions, and 3) established cytotoxicity-related genes^65,66^. The final list included the following genes: *UNC13D, NKG7, GZMB, TYROBP, HCST, KLRK1, NCR3, LILRB1, LAG3, INPP5D, IFNG, GZMH, FGFBP2, FURIN, IL18R1, GNLY, PRF1*.

### Cell-level linear mixed-effects models for cytotoxicity score analysis

To evaluate the association between DN2 identity and cytotoxic gene expression while accounting for donor-level variability, we applied cell-level linear mixed-effects models (LMM) using lme4 (v1.1-37) and lmerTest (v3.1-3). Two models were fitted. Model 1 tested whether DN2 status was associated with cytotoxicity score across all 36,538 B cells, with DN2 category (DN2-like vs non-DN2), disease status (SLE vs CTRL), age group (adult vs child), and log-transformed library size as fixed effects, and donor as a random intercept. The null model omitted the DN2 status term, and significance was assessed by likelihood ratio test. Model 2 tested whether disease status was associated with cytotoxicity score within DN2-like cells only (*n* = 7,498), using disease status, age group, and log-transformed library size as fixed effects with donor as a random intercept.

### K562 Killing assay

K562 cells were maintained in IMDM supplemented with 10% fetal bovine serum, 1% penicillin–streptomycin, and 1% L-glutamine at a density of 2 × 10⁵ cells/mL at 37 °C in a humidified incubator with 5% CO₂. Prior to each experiment, cells were collected, washed in PBS, and stained with 5 µM of Cell Trace Violet (Thermo Fisher, #C34571) for 10 minutes at 37 °C. Following incubation, cells were washed with five volumes of IMDM and resuspended in or t at a concentration of 2 × 10⁵ cells/mL. Cells were then allowed to rest for 10 minutes before plating 50 µL per well in a 96-well plate. Naïve B cells stimulated with the DN2-IC for 5 days or the DN2-IC and the rhIL-12A, or naïve B cells stimulated overnight with 10 ng/mL BAFF (BioLegend, #559602), were collected and washed in PBS for 15 minutes at 300 × g to enrich for viable cells. Cells were then plated at the indicated ratios with K562 target cells and incubated for 4 hours. Following co-culture, cells were collected, washed in PBS, and stained with Annexin V Apoptosis Detection Kits (eBioscience, #88-8005-74) according to the manufacturer’s instructions. Propidium iodide staining (25 µg/mL; BioLegend, #421301) was performed in FACS buffer within a 5-minute incubation at room temperature. Samples were acquired on a BD LSRFortessa™ flow cytometer (BD Biosciences). Instrument performance was verified using single-stained compensation beads. Data were collected using FACSDiva software (BD Biosciences) and analyzed with FlowJo software version 10.5.3 (BD Biosciences). The percentage of target cell killing was determined based on the proportion of dead cells identified within the Annexin V⁺/propidium iodide (PI)⁺ gate. Specific killing was calculated using the following formula: % Killing = [(percentage of dead cells in the presence of effectors − spontaneous death) × 100] / (100 − spontaneous death).

### CUT&RUN assay

Naive B cells (5–10 × 10^6) were washed twice with phosphate-buffered saline (PBS) and subjected to CUT&RUN with the CUT&LUNCH kit (#P-2035, EpiGentek) according to the manufacturer’s instructions, with minor modifications. Cells were incubated with either 2 µg of IRF4 antibody (Cell Signaling Technology, #4964) or a non-immune IgG control (provided with the kit) per reaction for 2 h at room temperature or overnight at 4°C. Following incubation, DNA was eluted and quantified using a Nanodrop spectrophotometer. For qPCR, 20–24 ng of eluted DNA was used per reaction. SYBR Green–based qPCR (Brilliant II SYBR® Green QPCR Master Mix #600828, Agilent) was performed using primers flanking a ∼100 bp region encompassing rs485499 following manufacture instructions. As a positive control for IRF4 binding, primers spanning the MYC super-enhancer were also included: Forward primer (rs485499 region): AAGATGGTGAGGAGGTTAG. Reverse primer (rs485499 region): TTTCTGCCCTCTCATAGTT. Forward primer (MYC super-enhancer): CCTCCTTATGCCTCTATCA. Reverse primer (MYC super-enhancer): GGAGAGTGGAGGAAAGAA. ΔCT values were calculated relative to the paired IgG control. For TaqMan genotyping, a predesigned SNP assay for rs485499 (assay ID:C 2423949_10, Life Technologies) was performed using TaqMan™ Genotyping Master Mix (#4371353, Life Technologies). Only heterozygous donors for rs485499 were included in this assay, and allele-specific signals were normalized to the corresponding IgG control. When IgG control samples were undetectable, IgG controls with comparable IRF4 signal were used as substitutes within the same experimental run. All donors, regardless of genotype, were included in the SYBR Green qPCR experiments.

### Statistical methods

Graphs and statistical analyses were performed using GraphPad Prism v10 (GraphPad Software). Paired two-tailed t-tests were used for comparisons between matched samples unless otherwise specified; one-tailed tests were used only where a directional hypothesis was defined a priori. Two-tailed Mann–Whitney tests were used for comparisons between independent groups. Differences were considered statistically significant at *p* ≤ 0.05. The Kruskal–Wallis test was used for comparisons involving more than two groups. Differences were considered statistically significant at *p* ≤ 0.05.

## Data and software availability

The single-cell RNA sequencing processed datasets and custom analysis scripts generated during this study are available in the GitHub repository from Gutierrez-Arcelus laboratory (https://github.com/gutierrez-arcelus-lab/IL12A_bcells). The raw sequencing data will be deposited in the Database of Genotypes and Phenotypes (dbGaP) under controlled access, and the accession number will be provided prior to publication.

## ACKNOWLEDGEMENTS

We thank Marina Sirota, and Taylor Kim for the help with rs485499 association studies. Yang Luo for help navigating the UK Biobank plasma proteome atlas. Ivan Zanoni, Marco Di Gioia, Davide Pallucci, Maggie Chang and Carola Vinuesa for thoughtful advice and critical comments. The Peter Nigrovic Laboratory for technical assistance and feedback. David Root and the Genetic Perturbation Platform (GPP) team at the Broad Institute for help with genome editing technologies. Tamara Salloum and the Center for Cellular Profiling at the Brigham and Women’s Hospital for technical support preparing single cell libraries.

Maria Gutierrez-Arcelus was supported by P30AR070253, the Lupus Research Alliance Lupus Innovation Award, the Lupus Research Alliance Empowering Research Career Development Award, the Vic Braden Family Arthritis National Research Foundation fellowship, the Gilead Sciences Rheumatology Research Scholars Award, the Boston Children’s Hospital Office of Faculty Development/Basic & Clinical Translational Research Executive Committees Faculty Career Development Fellowship. This work was supported by the National Institute of Allergy and Infectious Diseases grant 1R01AI198376, the Charles H. Hood Child Health Research Award, and the Cell Discovery Network, a collaborative initiative funded by the Manton Foundation and the Warren Alpert Foundation at Boston Children’s Hospital. Mariasilvia Colantuoni was supported by the Scientist Development Bridge Award from the Rheumatology Research Foundation. Joyce Chang was supported by the Rheumatology Research Foundation K Supplement and R Bridge Funding Awards and the Boston Children’s Hospital Office of Faculty Development/Basic & Clinical Translational Research Executive Committees Faculty Career Development Fellowship.

