## Extended Data for "Variant-to-function mapping in lupus links *IL12A* to the expansion of disease-associated B cells with a cytotoxic program"

**EXTENDED MATERIALS**

**EXTENDED TABLES**

**1. Extended Table 1.** Colocalization analysis between the chr3q25.33 SLE risk locus and *IL12A* eQTLs.

| **dataset_id** | **study_label** | **sample_group** | **tissue_label** | **condition_label** | **nsnps** | **PP.H4.abf** |
| --- | --- | --- | --- | --- | --- | --- |
| QTD000539 | TwinsUK | LCL | LCL | naive | 947 | 0.953 |
| QTD000110 | GEUVADIS | LCL | LCL | naive | 1123 | 0.94 |
| QTD000241 | GTEx | esophagus_mucosa | esophagus (mucosa) | naive | 1032 | 0.926 |
| QTD000171 | GTEx | brain_cortex | brain (cortex) | naive | 959 | 0.901 |
| QTD000156 | GTEx | brain_caudate | brain (caudate) | naive | 962 | 0.849 |
| QTD000075 | CommonMind | DLPFC_naive | brain (DLPFC) | naive | 1099 | 0.848 |
| QTD000579 | Walker_2019 | Neocortex | neocortex | naive | 1096 | 0.818 |
| QTD000211 | GTEx | breast | breast | naive | 1033 | 0.74 |
| QTD000373 | Lepik_2017 | blood | blood | naive | 886 | 0.548 |
| QTD000692 | Cytoimmgen | combined_CD4_Memory_STIM_5D | CD4+ memory T cell | anti-CD3-CD28_5D | 771 | 0.521 |
| QTD000146 | GTEx | brain_amygdala | brain (amygdala) | naive | 881 | 0.502 |
| QTD000221 | GTEx | LCL | LCL | naive | 976 | 0.456 |
| QTD000151 | GTEx | brain_anterior_cingulate_cortex | brain (anterior cingulate cortex) | naive | 901 | 0.443 |
| QTD000191 | GTEx | brain_nucleus_accumbens | brain (nucleus accumbens) | naive | 946 | 0.44 |
| QTD000716 | Cytoimmgen | TCM_STIM_5D | CD4+ T cell | anti-CD3-CD28_5D | 771 | 0.431 |
| QTD000095 | GENCORD | LCL | LCL | naive | 964 | 0.43 |
| QTD000181 | GTEx | brain_hippocampus | brain (hippocampus) | naive | 946 | 0.387 |
| QTD000316 | GTEx | skin_sun_exposed | skin | naive | 1043 | 0.343 |
| QTD000041 | Braineac2 | putamen | brain (putamen) | naive | 854 | 0.275 |
| QTD000366 | iPSCORE | iPSC | iPSC | naive | 862 | 0.258 |
| QTD000311 | GTEx | skin_not_sun_exposed | skin (suprapubic) | naive | 1045 | 0.194 |
| QTD000046 | Braineac2 | substantia_nigra | brain (substantia nigra) | naive | 648 | 0.187 |
| QTD000021 | BLUEPRINT | monocyte | monocyte | naive | 904 | 0.162 |
| QTD000186 | GTEx | brain_hypothalamus | brain (hypothalamus) | naive | 970 | 0.156 |
| QTD000196 | GTEx | brain_putamen | brain (putamen) | naive | 950 | 0.155 |
| QTD000296 | GTEx | pancreas | pancreas | naive | 1027 | 0.146 |
| QTD000306 | GTEx | prostate | prostate | naive | 1030 | 0.134 |
| QTD000166 | GTEx | brain_cerebellum | brain (cerebellum) | naive | 946 | 0.12 |
| QTD000286 | GTEx | nerve_tibial | tibial nerve | naive | 1033 | 0.119 |
| QTD000696 | Cytoimmgen | combined_CD4_Naive_STIM_5D | CD4+ T cell | anti-CD3-CD28_5D | 813 | 0.113 |
| QTD000634 | Jerber_2021 | Epen1_ROT_D52 | ependymal cell | rotenone_D52 | 887 | 0.104 |
| QTD000559 | Young_2019 | microglia_naive | microglia | naive | 869 | 0.103 |
| QTD000056 | CAP | LCL_naive | LCL | naive | 934 | 0.102 |
| QTD000479 | Schmiedel_2018 | CD4_T-cell_naive | CD4+ T cell | naive | 917 | 0.099 |
| QTD000564 | Aygun_2021 | Progenitor | neural progenitor | naive | 1004 | 0.099 |
| QTD000256 | GTEx | heart_left_ventricle | heart (left ventricle) | naive | 1024 | 0.096 |
| QTD000261 | GTEx | kidney_cortex | kidney (cortex) | naive | 815 | 0.096 |
| QTD000061 | CAP | LCL_statin | LCL | statin | 934 | 0.095 |
| QTD000036 | Bossini-Castillo_2019 | Treg_naive | Treg memory | naive | 866 | 0.092 |
| QTD000161 | GTEx | brain_cerebellar_hemisphere | brain (cerebellum) | naive | 930 | 0.091 |
| QTD000494 | Schmiedel_2018 | CD8_T-cell_anti-CD3-CD28 | CD8+ T cell | anti-CD3-CD28_4h | 917 | 0.09 |
| QTD000459 | Schmiedel_2018 | Th1-17_memory | Th17 cell | naive | 917 | 0.088 |
| QTD000301 | GTEx | pituitary | pituitary | naive | 987 | 0.086 |
| QTD000574 | PISA | pancreatic_islet | pancreatic islet | naive | 884 | 0.082 |
| QTD000326 | GTEx | spleen | spleen | naive | 1025 | 0.081 |
| QTD000524 | Steinberg_2020 | low_grade_cartilage_naive | cartilage | naive | 682 | 0.08 |
| QTD000276 | GTEx | minor_salivary_gland | minor salivary gland | naive | 977 | 0.077 |
| QTD000464 | Schmiedel_2018 | Treg_memory | Treg memory | naive | 917 | 0.076 |
| QTD000519 | Steinberg_2020 | synovium_naive | synovium | naive | 656 | 0.076 |
| QTD000016 | Alasoo_2018 | macrophage_IFNg+Salmonella | macrophage | IFNg_18h+Salmonella_5h | 797 | 0.074 |
| QTD000031 | BLUEPRINT | T-cell | CD4+ T cell | naive | 905 | 0.074 |
| QTD000011 | Alasoo_2018 | macrophage_Salmonella | macrophage | Salmonella_5h | 797 | 0.072 |
| QTD000504 | Schmiedel_2018 | monocyte_naive | monocyte | naive | 894 | 0.072 |
| QTD000691 | Cytoimmgen | combined_CD4_Memory_STIM_40H | CD4+ memory T cell | anti-CD3-CD28_40h | 769 | 0.072 |
| QTD000695 | Cytoimmgen | combined_CD4_Memory_STIM_16H | CD4+ memory T cell | anti-CD3-CD28_16h | 818 | 0.072 |
| QTD000701 | Cytoimmgen | TEM_STIM_5D | CD4+ T cell | anti-CD3-CD28_5D | 771 | 0.072 |
| QTD000544 | TwinsUK | skin | skin | naive | 951 | 0.07 |
| QTD000569 | Aygun_2021 | Neuron | neuron | naive | 989 | 0.07 |
| QTD000404 | PhLiPS | HLC | hepatocyte | naive | 949 | 0.069 |
| QTD000439 | Schmiedel_2018 | Tfh_memory | Tfh cell | naive | 917 | 0.068 |
| QTD000449 | Schmiedel_2018 | Th1_memory | Th1 cell | naive | 852 | 0.068 |
| QTD000051 | BrainSeq | brain | brain (DLPFC) | naive | 1148 | 0.067 |
| QTD000489 | Schmiedel_2018 | CD8_T-cell_naive | CD8+ T cell | naive | 917 | 0.067 |
| QTD000399 | PhLiPS | iPSC | iPSC | naive | 946 | 0.066 |
| QTD000529 | Steinberg_2020 | high_grade_cartilage_naive | cartilage | naive | 682 | 0.066 |
| QTD000601 | Perez_2022 | T8 | CD8+ T cell | naive | 975 | 0.066 |
| QTD000454 | Schmiedel_2018 | Th2_memory | Th2 cell | naive | 917 | 0.065 |
| QTD000499 | Schmiedel_2018 | monocyte_CD16_naive | CD16+ monocyte | naive | 919 | 0.065 |
| QTD000206 | GTEx | brain_substantia_nigra | brain (substantia nigra) | naive | 902 | 0.064 |
| QTD000346 | GTEx | uterus | uterus | naive | 917 | 0.064 |
| QTD000469 | Schmiedel_2018 | Treg_naive | Treg naive | naive | 917 | 0.064 |
| QTD000484 | Schmiedel_2018 | CD4_T-cell_anti-CD3-CD28 | CD4+ T cell | anti-CD3-CD28_4h | 917 | 0.064 |
| QTD000509 | Schmiedel_2018 | NK-cell_naive | NK cell | naive | 917 | 0.064 |
| QTD000176 | GTEx | brain_frontal_cortex | brain (DLPFC) | naive | 936 | 0.063 |
| QTD000379 | Nedelec_2016 | macrophage_Listeria | macrophage | Listeria_5h | 1072 | 0.063 |
| QTD000444 | Schmiedel_2018 | Th17_memory | Th17 cell | naive | 917 | 0.063 |
| QTD000006 | Alasoo_2018 | macrophage_IFNg | macrophage | IFNg_18h | 797 | 0.062 |
| QTD000126 | GTEx | adrenal_gland | adrenal gland | naive | 1024 | 0.062 |
| QTD000514 | Schwartzentruber_2018 | sensory_neuron | sensory neuron | naive | 851 | 0.062 |
| QTD000690 | Cytoimmgen | combined_CD4_Naive_STIM_40H | CD4+ T cell | anti-CD3-CD28_40h | 769 | 0.061 |
| QTD000693 | Cytoimmgen | combined_CD4_Naive_STIM_16H | CD4+ T cell | anti-CD3-CD28_16h | 773 | 0.06 |
| QTD000554 | van_de_Bunt_2015 | pancreatic_islet | pancreatic islet | naive | 831 | 0.058 |
| QTD000637 | Jerber_2021 | FPP_Naive_D11 | floor plate progenitor | naive_D11 | 885 | 0.058 |
| QTD000105 | GENCORD | T-cell | T cell | naive | 953 | 0.057 |
| QTD000414 | Quach_2016 | monocyte_LPS | monocyte | LPS_6h | 1139 | 0.057 |
| QTD000687 | Nathan_2022 | CD4+_Treg | CD4+ T cell | naive | 820 | 0.057 |
| QTD000351 | GTEx | vagina | vagina | naive | 925 | 0.056 |
| QTD000549 | TwinsUK | blood | blood | naive | 880 | 0.055 |
| QTD000136 | GTEx | artery_coronary | artery (coronary) | naive | 1022 | 0.054 |
| QTD000291 | GTEx | ovary | ovary | naive | 1004 | 0.054 |
| QTD000389 | Nedelec_2016 | macrophage_Salmonella | macrophage | Salmonella_5h | 1104 | 0.054 |
| QTD000597 | Perez_2022 | B | B cell | naive | 968 | 0.054 |
| QTD000201 | GTEx | brain_spinal_cord | brain (spinal cord) | naive | 906 | 0.053 |
| QTD000100 | GENCORD | fibroblast | fibroblast | naive | 970 | 0.052 |
| QTD000236 | GTEx | esophagus_gej | esophagus (gej) | naive | 1033 | 0.052 |
| QTD000321 | GTEx | small_intestine | small intestine | naive | 975 | 0.052 |
| QTD000141 | GTEx | artery_tibial | artery (tibial) | naive | 1041 | 0.051 |
| QTD000361 | HipSci | iPSC | iPSC | naive | 906 | 0.051 |
| QTD000429 | Quach_2016 | monocyte_IAV | monocyte | Influenza_6h | 1142 | 0.051 |
| QTD000336 | GTEx | testis | testis | naive | 1027 | 0.05 |
| QTD000424 | Quach_2016 | monocyte_R848 | monocyte | R848_6h | 1123 | 0.05 |
| QTD000331 | GTEx | stomach | stomach | naive | 1033 | 0.045 |
| QTD000226 | GTEx | colon_sigmoid | sigmoid colon | naive | 1034 | 0.044 |
| QTD000231 | GTEx | colon_transverse | transverse colon | naive | 1041 | 0.044 |
| QTD000341 | GTEx | thyroid | thyroid | naive | 1029 | 0.043 |
| QTD000116 | GTEx | adipose_subcutaneous | adipose | naive | 1036 | 0.041 |
| QTD000251 | GTEx | heart_atrial_appendage | heart (atrial appendage) | naive | 1027 | 0.041 |
| QTD000246 | GTEx | esophagus_muscularis | esophagus (muscularis) | naive | 1018 | 0.035 |
| QTD000121 | GTEx | adipose_visceral | adipose (visceral) | naive | 1033 | 0.033 |
| QTD000281 | GTEx | muscle | muscle | naive | 1027 | 0.03 |
| QTD000534 | TwinsUK | fat | adipose | naive | 955 | 0.029 |
| QTD000090 | FUSION | adipose_naive | adipose | naive | 905 | 0.025 |
| QTD000216 | GTEx | fibroblast | fibroblast | naive | 1034 | 0.021 |
| QTD000434 | ROSMAP | brain_naive | brain (DLPFC) | naive | 923 | 0.01 |
| QTD000474 | Schmiedel_2018 | B-cell_naive | B cell | naive | 894 | 0.002 |
| QTD000271 | GTEx | lung | lung | naive | 1033 | 0 |
| QTD000001 | Alasoo_2018 | macrophage_naive | macrophage | naive | NA | NA |
| QTD000026 | BLUEPRINT | neutrophil | neutrophil | naive | NA | NA |
| QTD000085 | FUSION | muscle_naive | muscle | naive | NA | NA |
| QTD000131 | GTEx | artery_aorta | artery (aorta) | naive | NA | NA |
| QTD000266 | GTEx | liver | liver | naive | NA | NA |
| QTD000356 | GTEx | blood | blood | naive | NA | NA |
| QTD000384 | Nedelec_2016 | macrophage_naive | macrophage | naive | NA | NA |
| QTD000394 | Peng_2018 | placenta_naive | placenta | naive | NA | NA |
| QTD000409 | Quach_2016 | monocyte_naive | monocyte | naive | NA | NA |
| QTD000419 | Quach_2016 | monocyte_Pam3CSK4 | monocyte | Pam3CSK4_6h | NA | NA |
| QTD000585 | Randolph_2021 | B_NI | B cell | naive | NA | NA |
| QTD000586 | Randolph_2021 | B_flu | B cell | Influenza_6h | NA | NA |
| QTD000587 | Randolph_2021 | CD4_T_NI | CD4+ T cell | naive | NA | NA |
| QTD000588 | Randolph_2021 | CD4_T_flu | CD4+ T cell | Influenza_6h | NA | NA |
| QTD000589 | Randolph_2021 | CD8_T_NI | CD8+ T cell | naive | NA | NA |
| QTD000590 | Randolph_2021 | CD8_T_flu | CD8+ T cell | Influenza_6h | NA | NA |
| QTD000591 | Randolph_2021 | NK_NI | NK cell | naive | NA | NA |
| QTD000592 | Randolph_2021 | NK_flu | NK cell | Influenza_6h | NA | NA |
| QTD000593 | Randolph_2021 | highly_infected_flu | monocyte | Influenza_6h | NA | NA |
| QTD000594 | Randolph_2021 | infected_monocytes_flu | monocyte | Influenza_6h | NA | NA |
| QTD000595 | Randolph_2021 | monocytes_NI | monocyte | naive | NA | NA |
| QTD000596 | Randolph_2021 | monocytes_flu | monocyte | Influenza_6h | NA | NA |
| QTD000598 | Perez_2022 | NK | NK cell | naive | NA | NA |
| QTD000599 | Perez_2022 | Prolif | NK cell | naive | NA | NA |
| QTD000600 | Perez_2022 | T4 | CD4+ T cell | naive | NA | NA |
| QTD000602 | Perez_2022 | cDC | dendritic cell | naive | NA | NA |
| QTD000603 | Perez_2022 | cM | monocyte | naive | NA | NA |
| QTD000604 | Perez_2022 | ncM | CD16+ monocyte | naive | NA | NA |
| QTD000605 | Perez_2022 | pDC | plasmacytoid dendritic cell | naive | NA | NA |
| QTD000606 | OneK1K | B_intermediate | B cell | naive | NA | NA |
| QTD000607 | OneK1K | B_memory | memory B cell | naive | NA | NA |
| QTD000608 | OneK1K | B_naive | B cell | naive | NA | NA |
| QTD000609 | OneK1K | CD14_Mono | monocyte | naive | NA | NA |
| QTD000610 | OneK1K | CD16_Mono | CD16+ monocyte | naive | NA | NA |
| QTD000611 | OneK1K | CD4_CTL | CD4+ CTL cell | naive | NA | NA |
| QTD000612 | OneK1K | CD4_Naive | CD4+ T cell | naive | NA | NA |
| QTD000613 | OneK1K | CD4_TCM | CD4+ TCM cell | naive | NA | NA |
| QTD000614 | OneK1K | CD4_TEM | CD4+ TEM cell | naive | NA | NA |
| QTD000615 | OneK1K | CD8_Naive | CD8+ T cell | naive | NA | NA |
| QTD000616 | OneK1K | CD8_TCM | CD8+ TCM cell | naive | NA | NA |
| QTD000617 | OneK1K | CD8_TEM | CD8+ TEM cell | naive | NA | NA |
| QTD000618 | OneK1K | HSPC | hematopoietic precursor cell | naive | NA | NA |
| QTD000619 | OneK1K | MAIT | MAIT cell | naive | NA | NA |
| QTD000620 | OneK1K | NK | NK cell | naive | NA | NA |
| QTD000621 | OneK1K | NK_CD56bright | CD56+ NK cell | naive | NA | NA |
| QTD000622 | OneK1K | NK_Proliferating | NK cell | naive | NA | NA |
| QTD000623 | OneK1K | Plasmablast | plasmablast | naive | NA | NA |
| QTD000624 | OneK1K | Platelet | platelet | naive | NA | NA |
| QTD000625 | OneK1K | Treg | Treg memory | naive | NA | NA |
| QTD000626 | OneK1K | cDC2 | dendritic cell | naive | NA | NA |
| QTD000627 | OneK1K | dnT | dnT cell | naive | NA | NA |
| QTD000628 | OneK1K | gdT | gdT cell | naive | NA | NA |
| QTD000629 | OneK1K | pDC | plasmacytoid dendritic cell | naive | NA | NA |
| QTD000630 | Jerber_2021 | NB_Naive_D11 | neuroblast | naive_D11 | NA | NA |
| QTD000631 | Jerber_2021 | U_Neur1_ROT_D52 | neuron | rotenone_D52 | NA | NA |
| QTD000632 | Jerber_2021 | P_FPP_ROT_D52 | floor plate progenitor | rotenone_D52 | NA | NA |
| QTD000633 | Jerber_2021 | DA_ROT_D52 | dopaminergic neuron | rotenone_D52 | NA | NA |
| QTD000635 | Jerber_2021 | P_FPP_Naive_D30 | floor plate progenitor | naive_D30 | NA | NA |
| QTD000636 | Jerber_2021 | U_Neur1_Naive_D30 | neuron | naive_D30 | NA | NA |
| QTD000638 | Jerber_2021 | FPP_Naive_D30 | floor plate progenitor | naive_D30 | NA | NA |
| QTD000639 | Jerber_2021 | P_Sert_Naive_D52 | serotonergic neuron | naive_D52 | NA | NA |
| QTD000640 | Jerber_2021 | Astro_Naive_D52 | astrocyte | naive_D52 | NA | NA |
| QTD000641 | Jerber_2021 | Epen2_Naive_D52 | ependymal cell | naive_D52 | NA | NA |
| QTD000642 | Jerber_2021 | FPP_Naive_D52 | floor plate progenitor | naive_D52 | NA | NA |
| QTD000643 | Jerber_2021 | Sert_Naive_D52 | serotonergic neuron | naive_D52 | NA | NA |
| QTD000644 | Jerber_2021 | P_FPP_Naive_D11 | floor plate progenitor | naive_D11 | NA | NA |
| QTD000645 | Jerber_2021 | Astro_ROT_D52 | astrocyte | rotenone_D52 | NA | NA |
| QTD000646 | Jerber_2021 | FPP_ROT_D52 | floor plate progenitor | rotenone_D52 | NA | NA |
| QTD000647 | Jerber_2021 | U_Neur3_ROT_D52 | neuron | rotenone_D52 | NA | NA |
| QTD000648 | Jerber_2021 | Sert_ROT_D52 | serotonergic neuron | rotenone_D52 | NA | NA |
| QTD000649 | Jerber_2021 | Epen1_Naive_D30 | ependymal cell | naive_D30 | NA | NA |
| QTD000650 | Jerber_2021 | P_FPP_Naive_D52 | floor plate progenitor | naive_D52 | NA | NA |
| QTD000651 | Jerber_2021 | Sert_Naive_D30 | serotonergic neuron | naive_D30 | NA | NA |
| QTD000652 | Jerber_2021 | U_Neur1_Naive_D52 | neuron | naive_D52 | NA | NA |
| QTD000653 | Jerber_2021 | Epen2_ROT_D52 | ependymal cell | rotenone_D52 | NA | NA |
| QTD000654 | Jerber_2021 | U_Neur2_Naive_D30 | neuron | naive_D30 | NA | NA |
| QTD000655 | Jerber_2021 | P_Sert_ROT_D52 | serotonergic neuron | rotenone_D52 | NA | NA |
| QTD000656 | Jerber_2021 | U_Neur3_Naive_D52 | neuron | naive_D52 | NA | NA |
| QTD000657 | Jerber_2021 | DA_Naive_D30 | dopaminergic neuron | naive_D30 | NA | NA |
| QTD000658 | Jerber_2021 | Epen1_Naive_D52 | ependymal cell | naive_D52 | NA | NA |
| QTD000659 | Jerber_2021 | DA_Naive_D52 | dopaminergic neuron | naive_D52 | NA | NA |
| QTD000660 | Nathan_2022 | CD4+_CCR4+ICOS+_central | CD4+ T cell | naive | NA | NA |
| QTD000661 | Nathan_2022 | CD4+_central | CD4+ T cell | naive | NA | NA |
| QTD000662 | Nathan_2022 | CD4+_CCR5+_cytotoxic | CD4+ T cell | naive | NA | NA |
| QTD000663 | Nathan_2022 | CD4+_CD161+_cytotoxic | CD4+ T cell | naive | NA | NA |
| QTD000664 | Nathan_2022 | CD8+_GZMK+ | CD4+ T cell | naive | NA | NA |
| QTD000665 | Nathan_2022 | CD8+_GZMB+ | CD4+ T cell | naive | NA | NA |
| QTD000666 | Nathan_2022 | CD4+_activated | CD4+ T cell | naive | NA | NA |
| QTD000667 | Nathan_2022 | Vd2 | CD4+ T cell | naive | NA | NA |
| QTD000668 | Nathan_2022 | CD4+_CD161+_Th2 | CD4+ T cell | naive | NA | NA |
| QTD000669 | Nathan_2022 | CD4-8+_PD-1+TIGIT+ | CD4+ T cell | naive | NA | NA |
| QTD000670 | Nathan_2022 | Vd1 | CD4+ T cell | naive | NA | NA |
| QTD000671 | Nathan_2022 | CD8+_central | CD4+ T cell | naive | NA | NA |
| QTD000672 | Nathan_2022 | CD4+_CD161+_Th1 | CD4+ T cell | naive | NA | NA |
| QTD000673 | Nathan_2022 | CD4+_Th17-1 | CD4+ T cell | naive | NA | NA |
| QTD000674 | Nathan_2022 | CD4+_HLA-DR+ | CD4+ T cell | naive | NA | NA |
| QTD000675 | Nathan_2022 | CD4+_CCR4+_central | CD4+ T cell | naive | NA | NA |
| QTD000676 | Nathan_2022 | CD8+_CXCR3+ | CD4+ T cell | naive | NA | NA |
| QTD000677 | Nathan_2022 | CD4+_Th2 | CD4+ T cell | naive | NA | NA |
| QTD000678 | Nathan_2022 | CD4+_CCR4+ | CD4+ T cell | naive | NA | NA |
| QTD000679 | Nathan_2022 | CD4+_CD38+ICOS+_central | CD4+ T cell | naive | NA | NA |
| QTD000680 | Nathan_2022 | CD4+_cytotoxic | CD4+ T cell | naive | NA | NA |
| QTD000681 | Nathan_2022 | CD4+_CD27+CD161+ | CD4+ T cell | naive | NA | NA |
| QTD000682 | Nathan_2022 | CD4+_RORC+_Treg | CD4+ T cell | naive | NA | NA |
| QTD000683 | Nathan_2022 | CD4+_Th1 | CD4+ T cell | naive | NA | NA |
| QTD000684 | Nathan_2022 | CD8+_activated | CD4+ T cell | naive | NA | NA |
| QTD000685 | Nathan_2022 | CD4+_CD27+ | CD4+ T cell | naive | NA | NA |
| QTD000686 | Nathan_2022 | CD4+_lncRNA | CD4+ T cell | naive | NA | NA |
| QTD000688 | Nathan_2022 | CD4+_Th17 | CD4+ T cell | naive | NA | NA |
| QTD000689 | Cytoimmgen | combined_CD4_Naive_UNS_16H | CD4+ T cell | naive | NA | NA |
| QTD000694 | Cytoimmgen | combined_CD4_Memory_UNS_16H | CD4+ memory T cell | naive | NA | NA |
| QTD000697 | Cytoimmgen | TN1_UNS_16H | CD4+ T cell | naive | NA | NA |
| QTD000698 | Cytoimmgen | TEM_STIM_16H | CD4+ T cell | anti-CD3-CD28_16h | NA | NA |
| QTD000699 | Cytoimmgen | TEMRA_STIM_5D | CD4+ T cell | anti-CD3-CD28_5D | NA | NA |
| QTD000700 | Cytoimmgen | TN_IFN_Lowly_Active_STIM_40H | CD4+ T cell | anti-CD3-CD28_40h | NA | NA |
| QTD000702 | Cytoimmgen | TN2_Lowly_Active_STIM_16H | CD4+ T cell | anti-CD3-CD28_16h | NA | NA |
| QTD000703 | Cytoimmgen | nTreg_STIM_40H | CD4+ T cell | anti-CD3-CD28_40h | NA | NA |
| QTD000704 | Cytoimmgen | TEM_STIM_40H | CD4+ T cell | anti-CD3-CD28_40h | NA | NA |
| QTD000705 | Cytoimmgen | TCM1_Lowly_Active_STIM_40H | CD4+ T cell | anti-CD3-CD28_40h | NA | NA |
| QTD000706 | Cytoimmgen | TN_STIM_5D | CD4+ T cell | anti-CD3-CD28_5D | NA | NA |
| QTD000707 | Cytoimmgen | TCM_UNS_16H | CD4+ T cell | naive | NA | NA |
| QTD000708 | Cytoimmgen | TCM_STIM_40H | CD4+ T cell | anti-CD3-CD28_40h | NA | NA |
| QTD000709 | Cytoimmgen | TEM_HLA+_STIM_40H | CD4+ T cell | anti-CD3-CD28_40h | NA | NA |
| QTD000710 | Cytoimmgen | TN_A_STIM_40H | CD4+ T cell | anti-CD3-CD28_40h | NA | NA |
| QTD000711 | Cytoimmgen | TN_HSP_STIM_5D | CD4+ T cell | anti-CD3-CD28_5D | NA | NA |
| QTD000712 | Cytoimmgen | TN_A_STIM_16H | CD4+ T cell | anti-CD3-CD28_16h | NA | NA |
| QTD000713 | Cytoimmgen | T_NFKB_STIM_16H | CD4+ T cell | anti-CD3-CD28_16h | NA | NA |
| QTD000714 | Cytoimmgen | TN3_UNS_16H | CD4+ T cell | naive | NA | NA |
| QTD000715 | Cytoimmgen | TCM1_Lowly_Active_STIM_16H | CD4+ T cell | anti-CD3-CD28_16h | NA | NA |
| QTD000717 | Cytoimmgen | TN_NFKB_STIM_5D | CD4+ T cell | anti-CD3-CD28_5D | NA | NA |
| QTD000718 | Cytoimmgen | TN_B_STIM_16H | CD4+ T cell | anti-CD3-CD28_16h | NA | NA |
| QTD000719 | Cytoimmgen | TEMRA_UNS_16H | CD4+ T cell | naive | NA | NA |
| QTD000720 | Cytoimmgen | TN2_UNS_16H | CD4+ T cell | naive | NA | NA |
| QTD000721 | Cytoimmgen | nTreg_STIM_16H | CD4+ T cell | anti-CD3-CD28_16h | NA | NA |
| QTD000722 | Cytoimmgen | TN1_Lowly_Active_STIM_40H | CD4+ T cell | anti-CD3-CD28_40h | NA | NA |
| QTD000723 | Cytoimmgen | TN2_STIM_16H | CD4+ T cell | anti-CD3-CD28_16h | NA | NA |
| QTD000724 | Cytoimmgen | TEM_Lowly_Active_STIM_40H | CD4+ T cell | anti-CD3-CD28_40h | NA | NA |
| QTD000725 | Cytoimmgen | TN3_Lowly_Active_STIM_16H | CD4+ T cell | anti-CD3-CD28_16h | NA | NA |
| QTD000726 | Cytoimmgen | TCM_STIM_16H | CD4+ T cell | anti-CD3-CD28_16h | NA | NA |
| QTD000727 | Cytoimmgen | TN_cycling_STIM_5D | CD4+ T cell | anti-CD3-CD28_5D | NA | NA |
| QTD000728 | Cytoimmgen | TN_IFN_STIM_5D | CD4+ T cell | anti-CD3-CD28_5D | NA | NA |
| QTD000729 | Cytoimmgen | TM_cycling_STIM_5D | CD4+ T cell | anti-CD3-CD28_5D | NA | NA |
| QTD000730 | Cytoimmgen | TEM_HLA+_STIM_5D | CD4+ T cell | anti-CD3-CD28_5D | NA | NA |
| QTD000731 | Cytoimmgen | TEMRA_STIM_16H | CD4+ T cell | anti-CD3-CD28_16h | NA | NA |
| QTD000732 | Cytoimmgen | TM_ER-stress_STIM_40H | CD4+ T cell | anti-CD3-CD28_40h | NA | NA |
| QTD000733 | Cytoimmgen | TN1_STIM_40H | CD4+ T cell | anti-CD3-CD28_40h | NA | NA |
| QTD000734 | Cytoimmgen | TEM_UNS_16H | CD4+ T cell | naive | NA | NA |
| QTD000735 | Cytoimmgen | TN2_STIM_40H | CD4+ T cell | anti-CD3-CD28_40h | NA | NA |
| QTD000736 | Cytoimmgen | TEMRA_Lowly_Active_STIM_16H | CD4+ T cell | anti-CD3-CD28_16h | NA | NA |
| QTD000737 | Cytoimmgen | TN_IFN_STIM_40H | CD4+ T cell | anti-CD3-CD28_40h | NA | NA |
| QTD000738 | Cytoimmgen | TN_cycling_STIM_40H | CD4+ T cell | anti-CD3-CD28_40h | NA | NA |
| QTD000739 | Cytoimmgen | TN_NFKB_STIM_40H | CD4+ T cell | anti-CD3-CD28_40h | NA | NA |
| QTD000740 | Cytoimmgen | TCM2_Lowly_Active_STIM_16H | CD4+ T cell | anti-CD3-CD28_16h | NA | NA |
| QTD000741 | Cytoimmgen | TN_C_STIM_16H | CD4+ T cell | anti-CD3-CD28_16h | NA | NA |
| QTD000742 | Cytoimmgen | TEM_Lowly_Active_STIM_16H | CD4+ T cell | anti-CD3-CD28_16h | NA | NA |
| QTD000743 | Cytoimmgen | T_ER-stress_STIM_5D | CD4+ T cell | anti-CD3-CD28_5D | NA | NA |
| QTD000744 | Cytoimmgen | TCM2_Lowly_Active_STIM_40H | CD4+ T cell | anti-CD3-CD28_40h | NA | NA |
| QTD000745 | Cytoimmgen | TN_IFN_STIM_16H | CD4+ T cell | anti-CD3-CD28_16h | NA | NA |
| QTD000746 | Cytoimmgen | HSP_STIM_16H | CD4+ T cell | anti-CD3-CD28_16h | NA | NA |
| QTD000747 | Cytoimmgen | nTreg_UNS_16H | CD4+ T cell | naive | NA | NA |
| QTD000748 | Cytoimmgen | TN_IFN_Lowly_Active_STIM_16H | CD4+ T cell | anti-CD3-CD28_16h | NA | NA |
| QTD000749 | Cytoimmgen | TN-TCM_CXCR4+_STIM_40H | CD4+ T cell | anti-CD3-CD28_40h | NA | NA |
| QTD000750 | Cytoimmgen | TEMRA_STIM_40H | CD4+ T cell | anti-CD3-CD28_40h | NA | NA |
| QTD000751 | Cytoimmgen | TN1_Lowly_Active_STIM_16H | CD4+ T cell | anti-CD3-CD28_16h | NA | NA |

NA = Not available; data for the the 3q25.33 SLE locus were not found in those datasets.

**2. Extended Table 2.** Putative enhancer-associated variants located within active or poised regulatory elements.

| **rsID** | **enhancer** | **DNase I** | **H3K4me1** | **H3K4me3** | **H3K27ac** | **ATAC-seq** | **GRO-seq** |
| --- | --- | --- | --- | --- | --- | --- | --- |
| rs529951838 | 1 | 1 | 0 | 0 | 1 | 1 | 0 |
| rs629209 | 1 | 1 | 0 | 1 | 0 | 1 | 1 |
| rs564799 | 1 | 0 | 1 | 0 | 1 | 0 | 0 |
| rs564976 | 1 | 0 | 1 | 0 | 1 | 1 | 0 |
| rs138653054 | 1 | 0 | 1 | 0 | 1 | 1 | 0 |
| rs480134 | 1 | 0 | 1 | 0 | 1 | 0 | 0 |
| rs586094 | 1 | 0 | 1 | 0 | 1 | 0 | 0 |
| rs480913 | 1 | 0 | 1 | 0 | 1 | 0 | 0 |
| rs1874886 | 1 | 0 | 1 | 0 | 1 | 0 | 0 |
| rs587422 | 1 | 0 | 1 | 0 | 1 | 1 | 0 |
| rs483714 | 1 | 0 | 1 | 0 | 1 | 1 | 0 |
| rs10575904 | 1 | 0 | 1 | 0 | 1 | 1 | 0 |
| rs484600 | 1 | 0 | 0 | 0 | 1 | 1 | 0 |
| rs485789 | 1 | 1 | 0 | 0 | 1 | 1 | 0 |
| rs600519 | 1 | 0 | 1 | 1 | 1 | 1 | 0 |
| rs6808498 | 1 | 0 | 1 | 1 | 1 | 0 | 0 |
| rs6771363 | 1 | 0 | 1 | 1 | 1 | 0 | 0 |
| rs6808518 | 1 | 0 | 1 | 1 | 1 | 0 | 0 |
| rs2936303 | 1 | 0 | 1 | 0 | 0 | 0 | 0 |
| rs2936302 | 1 | 0 | 0 | 0 | 0 | 0 | 0 |
| rs7640862 | 1 | 0 | 0 | 0 | 0 | 0 | 0 |
| rs2914116 | 1 | 0 | 0 | 0 | 0 | 0 | 0 |
| rs9845010 | 1 | 0 | 0 | 0 | 0 | 0 | 0 |
| rs202034560 | 1 | 0 | 0 | 0 | 0 | 0 | 0 |
| rs669003 | 1 | 0 | 0 | 0 | 0 | 0 | 0 |
| rs545143 | 1 | 0 | 0 | 0 | 0 | 0 | 0 |
| rs545232 | 1 | 0 | 0 | 0 | 0 | 0 | 0 |
| rs547875 | 1 | 0 | 0 | 0 | 0 | 0 | 0 |
| rs548018 | 1 | 0 | 0 | 0 | 0 | 0 | 0 |
| rs1651081 | 1 | 0 | 0 | 0 | 0 | 0 | 0 |
| rs571099 | 1 | 0 | 0 | 0 | 0 | 0 | 0 |
| rs11383537 | 1 | 0 | 0 | 0 | 0 | 0 | 0 |
| rs522127 | 1 | 0 | 0 | 0 | 0 | 0 | 0 |
| rs523886 | 1 | 1 | 0 | 0 | 0 | 0 | 0 |
| rs485499 | 1 | 1 | 1 | 0 | 1 | 1 | 1 |
| rs2647928 | 1 | 0 | 0 | 0 | 0 | 0 | 0 |

Variants were annotated in a binary manner: 1 if the variant falls within a region defined by the given chromatin mark, and 0 otherwise.

**3. Extended Table 3.** Tomtom analysis of transcription factors binding motifs

| **Query ID** | **Target ID** | **Optimaloffset** | **p-value** | **E-value** | **q-value** | **Overlap** | **Query consensus** | **Target consensus** | **Orientation** |
| --- | --- | --- | --- | --- | --- | --- | --- | --- | --- |
| GGAMATGA | STAT2_HUMAN.H11MO.0.A | 2 | 0.00094176 | 0.377647 | 0.337947 | 8 | GGAAATGA | AGGAAAATGAAACTGAAAG | + |
| GGAMATGA | ETV5_HUMAN.H11MO.0.C | 5 | 0.00097987 | 0.392928 | 0.337947 | 8 | GGAAATGA | GAGCAGGAAGTGAG | + |
| GGAMATGA | BC11A_HUMAN.H11MO.0.A | 6 | 0.00131371 | 0.526798 | 0.337947 | 8 | GGAAATGA | AAAAGAGGAAGTGAAAA | + |
| GGAMATGA | ZN816_HUMAN.H11MO.0.C | 9 | 0.00207663 | 0.83273 | 0.353793 | 8 | GGAAATGA | AAAAAAGGGGGACATGCAGGG | + |
| GGAMATGA | STAT3_HUMAN.H11MO.0.A | 5 | 0.00235783 | 0.945489 | 0.353793 | 7 | GGAAATGA | TTCCGGGAAATG | - |
| GGAMATGA | IRF8_HUMAN.H11MO.0.B | 6 | 0.00333678 | 1.33805 | 0.353793 | 8 | GGAAATGA | AAAAGAGGAAGTGAAAGTAA | + |
| GGAMATGA | HIF1A_HUMAN.H11MO.0.C | 0 | 0.00338901 | 1.35899 | 0.353793 | 8 | GGAAATGA | GGACGTGC | + |
| GGAMATGA | SPIB_HUMAN.H11MO.0.A | 7 | 0.00373972 | 1.49963 | 0.353793 | 8 | GGAAATGA | AAAAAGAGGAAGTGAAA | + |
| GGAMATGA | ELK1_HUMAN.H11MO.0.B | 3 | 0.00464151 | 1.86125 | 0.353793 | 8 | GGAAATGA | ACCGGAAGTGG | + |
| GGAMATGA | SPI1_HUMAN.H11MO.0.A | 7 | 0.00490609 | 1.96734 | 0.353793 | 8 | GGAAATGA | AAAAAGAGGAAGTGAAA | + |
| GGAMATGA | IRF4_HUMAN.H11MO.0.A | 6 | 0.00504279 | 2.02216 | 0.353793 | 8 | GGAAATGA | AAAAGAGGAAGTGAAACT | + |
| GGAMATGA | ELF2_HUMAN.H11MO.0.C | 5 | 0.00777803 | 3.11899 | 0.450855 | 8 | GGAAATGA | AACCCGGAAGTGG | + |
| GGAMATGA | STAT1_HUMAN.H11MO.0.A | 10 | 0.00932732 | 3.74026 | 0.450855 | 8 | GGAAATGA | CCCCTTTCCTGGAAATCAC | + |
| GGAMATGA | ELK4_HUMAN.H11MO.0.A | 4 | 0.0100978 | 4.04922 | 0.450855 | 8 | GGAAATGA | GACCGGAAGTGG | + |
| GGAMATGA | GABPA_HUMAN.H11MO.0.A | 6 | 0.0102739 | 4.11985 | 0.450855 | 8 | GGAAATGA | GGAACCGGAAGTGG | + |
| GGAMATGA | REL_HUMAN.H11MO.0.B | 5 | 0.0106867 | 4.28535 | 0.450855 | 8 | GGAAATGA | TGAAGGGAAATTCCA | + |
| GGAMATGA | ELF5_HUMAN.H11MO.0.A | 7 | 0.0116652 | 4.67773 | 0.450855 | 8 | GGAAATGA | GGAAGGAGGAAGTGG | + |
| GGAMATGA | IRF1_HUMAN.H11MO.0.A | 0 | 0.0129511 | 5.19338 | 0.450855 | 8 | GGAAATGA | GAAAATGAAAGTGAAAGTAA | + |
| GGAMATGA | ERG_HUMAN.H11MO.0.A | 5 | 0.01303 | 5.22503 | 0.450855 | 8 | GGAAATGA | GGGCAGGAAGTGG | + |
| GGAMATGA | ELF3_HUMAN.H11MO.0.A | 6 | 0.0132668 | 5.31999 | 0.450855 | 8 | GGAAATGA | GAACCAGGAAGTGG | + |
| GGAMATGA | LEF1_HUMAN.H11MO.0.A | 1 | 0.0144356 | 5.78867 | 0.450855 | 8 | GGAAATGA | AGCAAATCAAAGGA | - |
| GGAMATGA | HXC9_HUMAN.H11MO.0.C | 0 | 0.0146173 | 5.86152 | 0.450855 | 8 | GGAAATGA | GGCAATAAAA | - |
| GGAMATGA | ZN528_HUMAN.H11MO.0.C | 2 | 0.0148976 | 5.97395 | 0.450855 | 8 | GGAAATGA | TCAGAAATGGCTTCCCTGGG | - |
| GGAMATGA | Z354A_HUMAN.H11MO.0.C | 9 | 0.0152534 | 6.11661 | 0.450855 | 8 | GGAAATGA | ACATTAAATGTAAATGGACTAAAT | - |
| GGAMATGA | EHF_HUMAN.H11MO.0.B | 6 | 0.0157035 | 6.29712 | 0.450855 | 8 | GGAAATGA | GAACCAGGAAGTGGC | + |
| GGAMATGA | P73_HUMAN.H11MO.0.A | 9 | 0.0157736 | 6.3252 | 0.450855 | 8 | GGAAATGA | GGCATGTCTGGGCATGTCT | - |
| GGAMATGA | ZN680_HUMAN.H11MO.0.C | 13 | 0.0170412 | 6.83352 | 0.45379 | 7 | GGAAATGA | CCTCATTCTTCTTGGACATG | - |
| GGAMATGA | NF2L1_HUMAN.H11MO.0.C | -3 | 0.0171781 | 6.88842 | 0.45379 | 5 | GGAAATGA | AATGACT | - |
| GGAMATGA | ETS1_HUMAN.H11MO.0.A | 0 | 0.0176403 | 7.07376 | 0.45379 | 8 | GGAAATGA | AGACAGGAAGTGG | + |
| GGAMATGA | ZN350_HUMAN.H11MO.0.C | 2 | 0.0187639 | 7.52432 | 0.46003 | 8 | GGAAATGA | TAGGTCATAAAAGGACTG | - |
| GGAMATGA | BHE40_HUMAN.H11MO.0.A | 1 | 0.019075 | 7.64909 | 0.46003 | 8 | GGAAATGA | GGCACGTGAC | + |
| GGAMATGA | P53_HUMAN.H11MO.0.A | 0 | 0.0202515 | 8.12085 | 0.473602 | 8 | GGAAATGA | GGGCATGTCTGGGCATGTCT | - |
| GGAMATGA | P63_HUMAN.H11MO.0.A | 10 | 0.0212733 | 8.53058 | 0.477363 | 8 | GGAAATGA | GGGCATGCCTGGACATGCC | - |
| GGAMATGA | ETV2_HUMAN.H11MO.0.B | 7 | 0.0216983 | 8.70101 | 0.477363 | 8 | GGAAATGA | GGAAACAGGAAGTGGG | + |
| GGAMATGA | PRDM1_HUMAN.H11MO.0.A | 0 | 0.022268 | 8.92946 | 0.477363 | 8 | GGAAATGA | GAAAGTGAAAGTGA | + |
| GGAMATGA | ETV1_HUMAN.H11MO.0.A | 4 | 0.0242927 | 9.74136 | 0.493358 | 7 | GGAAATGA | GGCCGGAAGTG | + |
| GGAMATGA | ETV4_HUMAN.H11MO.0.B | 4 | 0.0242927 | 9.74136 | 0.493358 | 7 | GGAAATGA | AACAGGAAGGG | + |

Tomtom (Motif Comparison Tool): Version 5.5.7 compiled on Aug 29 2024 at 17:50:26

The format of this file is described at <https://meme-suite.org/meme/doc/tomtom-output-format.html>.

Tomtom -no-ssc -oc . -verbosity 1 -min-overlap 5 -mi 1 -dist pearson -evalue -thresh 10.0 -time 300 motif_number_4_from_meme.html.txt db/HUMAN/HOCOMOCOv11_core_HUMAN_mono_meme_format.meme

**4. Extended Table 4.** Primers sequences for quantitative PCR.

| **Target gene** | **Forward** | **Reverse** | **Supplier** |
| --- | --- | --- | --- |
| *GAPDH* | CCACATCGCTCAGACACCAT | GGCAACAATATCCACTTTACCAGAGT | Origene |
|  |  |  | #HP205798 |
| *RPL10A* | CCACATCGCTCAGACACCAT | GACACCAAGAAGTTGACAGCCAG | Origene |
|  |  |  | #HP209896 |
| *IL12A* | TGCCTTCACCACTCCCAAAACC | CAATCTCTTCAGAAGTGCAAGGG | Origene |
|  |  |  | #HP200821 |
| *IL12B* | GACATTCTGCGTTCAGGTCCAG | CATTTTTGCGGCAGATGACCGTG | Origene |
|  |  |  | #HP205923 |
| *EBI3* | CTGGATCCGTTACAAGCGTCAG | CACTTGGACGTAGTACCTGGCT | Origene |
|  |  |  | #HP208836 |
| *IRF4* | GAACGAGGAGAAGAGCATCTTCC | CGATGCCTTCTCGGAACTTTCC | Origene |
|  |  |  | #HP206140 |

**5. Extended Table 5.** Antibodies used for the immunophenotyping of B cells.

| **Antibody** | **Fluorochrome** | **Cat. Number** | **Clone** | **Supplier** |
| --- | --- | --- | --- | --- |
| CD19 | BV687 | 302240 | HIB19 | BioLegend |
| CD27 | BV605 | 562655 | L128 | BD Biosciences |
| IGD | PerCP Cyanine5 5 | 348208 | IA6-2 | BioLegend |
| CD21 | FITC | 354909 | Bu32 | BioLegend |
| CD11c | PE | 555392 | B-ly6 | BD Biosciences |
| CD38 | BV421 | 303525 | HIT2 | BioLegend |
| CD138 | PE-Texas red | 356529 | MI15 | BioLegend |
| LAG3 | BV421 | 369313 | 11C3C65 | BioLegend |
| IL18R1 | APC | 313813 | H44 | BioLegend |
| GZMB | PE-Texas red | 562462 | GB11 | BD Biosciences |
| GNLY | PE-Cy7 | 348011 | DH2 | BioLegend |
| PRF1 | APC | 154303 | S16009A | BioLegend |
| Mouse IgG1, κ Isotype Control | PE-CF594 | 562292 | MOPC-21 | BioLegend |
| Mouse IgG1 κ Isotype Control | PE-Cy7 | 557872 | MOPC-21 | BD Biosciences |
| Rat IgG2a, κ Isotype Control | APC | 400511 | RTK2758 | Legend |

**6. Extended Table 6.** Gating strategies for identifying distinct cell populations in PBMCs and B cells.

| **Population** | **Markers** |
| --- | --- |
| Naïve | CD19+ CD27- **IgD+** |
| Switched Memory | CD19+ **CD27+ IgD-** |
| Unswitched Memory | CD19+ **CD27+ IgD+** |
| Double negative | CD19+ **CD27- IgD-** |
| Double negative 1 (DN1) | CD19+ CD27- IgD- **CD21+** |
| Double negative 2 (DN2) | CD19+ CD27- IgD- **CD21-** **CD11c+** |
| Double negative 3 (DN3) | CD19+ CD27- IgD- **CD21- CD11c-** |
| Double negative 4 (DN4) | CD19+ CD27- IgD- **CD21+ CD11c+** |
| Memory CD11c+ | CD19+ **CD27+ IgD- CD11c+** |
| Plasma blasts | CD19+ CD27+ IgD- **CD38+** |
| Plasma cells | CD19+ CD27+ IgD- CD38+ **CD138+** |
| IL18R1+ B cells | CD19+ CD27- IgD- CD21- CD11c- **IL18R1+** |
| LAG3+ B cells | CD19+ CD27- IgD- CD21- CD11c- **LAG3+** |
| IL18R1+ DN2 cells | CD19+ CD27- IgD- **CD21- CD11c+ IL18R1+** |
| LAG3+ DN2 cells | CD19+ CD27- IgD- **CD21- CD11c+ LAG3+** |
| GZMB+ B cells | CD19+ **GZMB+** |
| GZMB+ Cd11C+ B cells | CD19+ **CD11C+ GZMB+** |
| PRF1+ B cells | CD19+ **PRF1+** |
| PRF1+ Cd11C+ B cells | CD19+ **CD11C+PRF1+** |
| GNLY+ B cells | CD19+ **GNLY+** |
| GNLY+ Cd11C+ B cells | CD19+ **CD11C+GNLY+** |
| NKG7+ B cells | CD19+ **NKG7+** |
| NKG7+ Cd11C+ B cells | CD19+ **CD11C+NKG7+** |
| K562 Annexin V+PI+ | CTV+ **Annexin V+PI+** |
| K562 Annexin V+PI- | CTV+ **Annexin V+PI-** |
| K562 Annexin V-PI+ | CTV+ **Annexin V-PI+** |

The surface markers used to identify the cell subsets are in bold.

**7. Extended Table 7.** sgRNA sequences for CRISPR experiments.

| **Target gene/region** | **Sequence** |
| --- | --- |
| *IL12A* | CCAGGGUAGCCACAAGGAGG |
|  | GGUCUGGAGUGGCCACGGGG |
|  | UGACGGCCCUCAGCAGGUUU |
| *EBI3* | CACCCUGUGCAGGCUCGGCA |
|  | GGAUGUCCAGCUGUUCUCCA |
|  | AGCUGCUGCUGGAGCCCCAG |
| *IRF4* | CACGCGGGGCAUGAACCUGG |
|  | GCGCGGUGAGCUGCGGCAAC |
|  | AGAGCAUCUUCCGCAUCCCC |
| Scramble | GCACUACCAGAGCUAACUCA |
| rs485499-containing region | AACACTGTGAAACTATGAGA |
| rs564965-containing region | GTGGGGATGCTTCTAATAGT |
| rs564965_ABE8e | CTGGGACATGAGTTTACCGT |

**8. Extended Table 8.** Human TOTAL-Seq C Repertoire (5') HASHING Abs

| **Hashtag** | **Cat #** | **Sample** |
| --- | --- | --- |
| #1 | 394661 | HD1 UTC |
| #2 | 394663 | HD2 UTC |
| #3 | 394665 | HD3 UTC |
| #4 | 394667 | HD4 UTC |
| #5 | 394669 | HD5 UTC |
| #6 | 394671 | HD1 rhIL-12A |
| #7 | 394673 | HD2 rhIL-12A |
| #8 | 394675 | HD3 rhIL-12A |
| #9 | 394677 | HD4 rhIL-12A |
| #10 | 394679 | HD5 rhIL-12A |

**EXTENDED METHODS:**

**Immunoblot analysis**

Proteins were extracted using ice-cold RIPA Lysis and Extraction Buffer (#PI89900, Thermo Scientific) supplemented with 100X Halt Protease Inhibitor Cocktail (#PI78429, Thermo Scientific). Lysates were quantified using the Pierce BCA Protein Assay Kit (#23227, Thermo Scientific), and 5–50 µg of protein were loaded onto 4–20% SDS-polyacrylamide gels (Bio-Rad, #4561094). Following electrophoresis, analytes were wet transferred to a PVDF membrane using a Mini Trans-Blot Cell (Bio-Rad) apparatus at 200 mA for 1.5 hours. Membranes were blocked for 2–4 hours (5% w/v BSA, 1X TBS, 0.1% Tween 20) and incubated overnight at 4°C with a primary antibody against IRF4 (#4964, Cell Signaling Technology, 1:1000). The following day, membranes were washed four times with 1X TBS-0.2% Tween 20 and incubated with HRP-conjugated Goat anti-Rabbit IgG secondary antibody (#31460, Thermo Scientific, 1:1000) for 1 hour at room temperature. After four additional washes, bands were detected using SuperSignal West Femto Maximum Sensitivity Substrate (#34095, Thermo Scientific) and imaged with an iBright Imaging System (Invitrogen). β-actin (Mouse monoclonal Anti-β-Actin-Peroxidase, 1:25000, 1 hour RT incubation) was used as a loading control.

**Transcription factor binding motif analysis**

A ~200 bp region surrounding the rs485499 variant was analyzed using the Motif Discovery tool in the MEME Suite^1^ to identify overlapping binding motifs. The identified motif flanking rs485499 (GGAA/CATGA) was then queried against the human motif database using Tomtom^1^. IRF4 was identified as a top candidate, ranking within the top 37 targets (*p* = 0.0050).

**EXTENDED DATA FIGURES.**

**Extended Figure 1.**

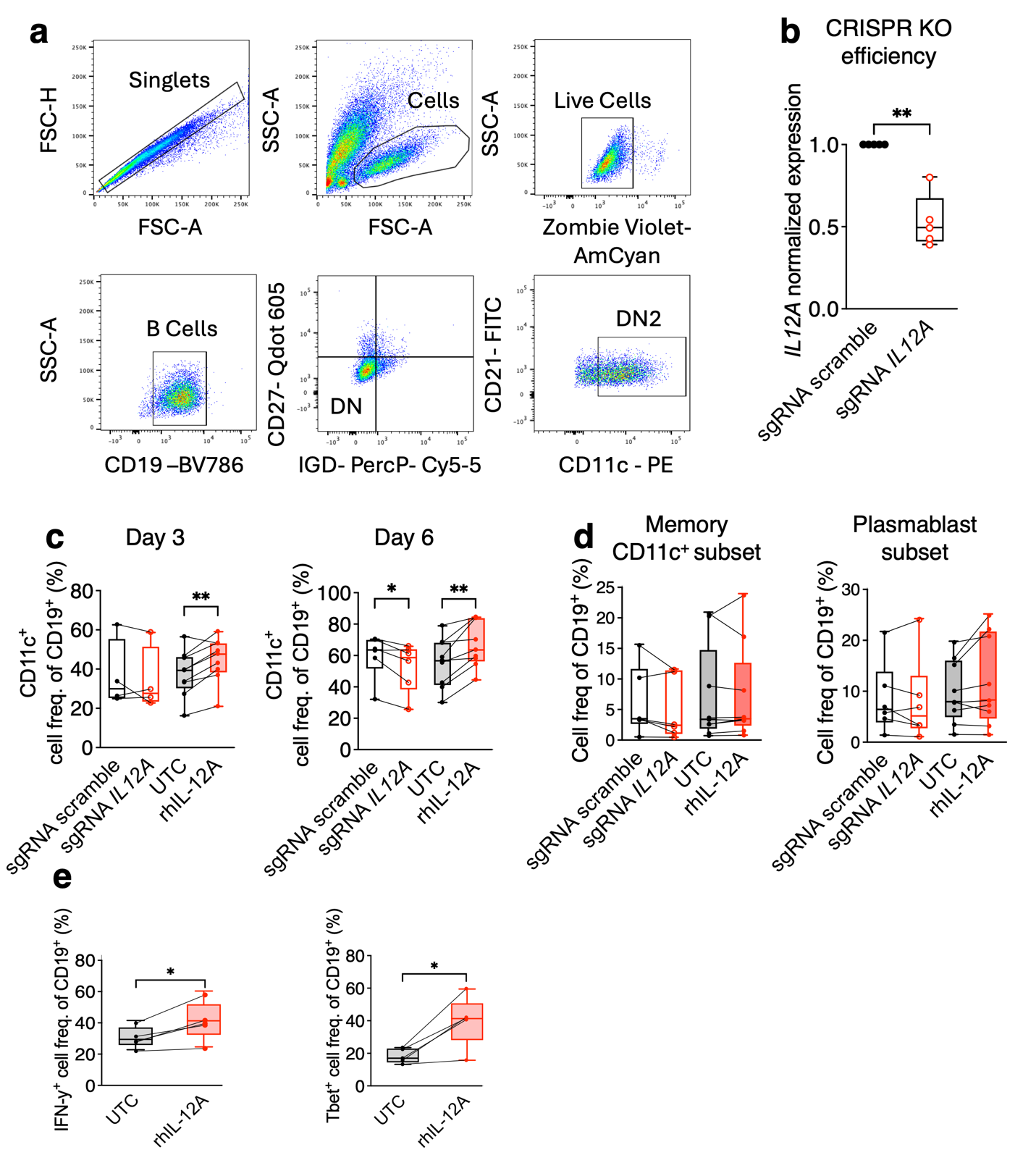

**Extended Figure 1. IL-12A promotes DN2 B cell differentiation**

**(a)** Representative gating strategy for FACS-based immunophenotyping of DN2 B cells (CD19⁺ IgD⁻ CD27⁻ CD21⁻ CD11c⁺). Gating is shown for naïve B cells after 6 days of *in vitro* differentiation with the DN2-IC. **(b)** *IL12A* CRISPR KO efficiency, measured by *IL12A* gene expression at day 3–4 post-stimulation in naïve B cells treated with DN2-IC. sgRNA scramble: cells nucleofected with the scramble sgRNA at 1-hour post-activation. sgRNA *IL12A*: cells nucleofected with sgRNA targeting *IL12A* gene at 1-hour post-activation. **(c)** Frequency of total CD11c^+^ B cells at day 3 and day 6 following *IL12A* CRISPR KO or rhIL-12A supplementation. **(d)** Frequencies of memory CD11c^+^ and plasmablast subsets at day 6 following *IL12A* CRISPR KO or rhIL-12A treatment. The gating strategies used for defining these populations are reported in Extended Table 6. **(e)** Percentages of IFNy^+^ and Tbet^+^ B cells at day 3 post-DN2-IC stimulation in the presence (rhIL-12A) or absence (UTC) of recombinant IL-12A. Data are presented as box plots showing the median and 25th/75th percentiles; whiskers indicate the range. Individual data points represent biological replicates (donors, n = 4 to 9). Cell populations are calculated as percentages of total CD19^+^ B cells. sgRNA scramble: cells nucleofected with the scramble sgRNA. sgRNA *IL12A*: cells nucleofected with sgRNA targeting *IL12A* gene. UTC: untreated control, naive B cells stimulated with the DN2-IC. RhIL-12A: naive B cells stimulated with the DN2-IC in the presence of rhIL12A (200ng/ml). **(b)** Gene expression is normalized to *GAPDH* and expressed relative to the scrambled (sgRNA scramble) control using the ΔΔCt method. Two-tailed paired t-test: **p* < 0.05, ***p* < 0.01.

**Extended Figure 2.**

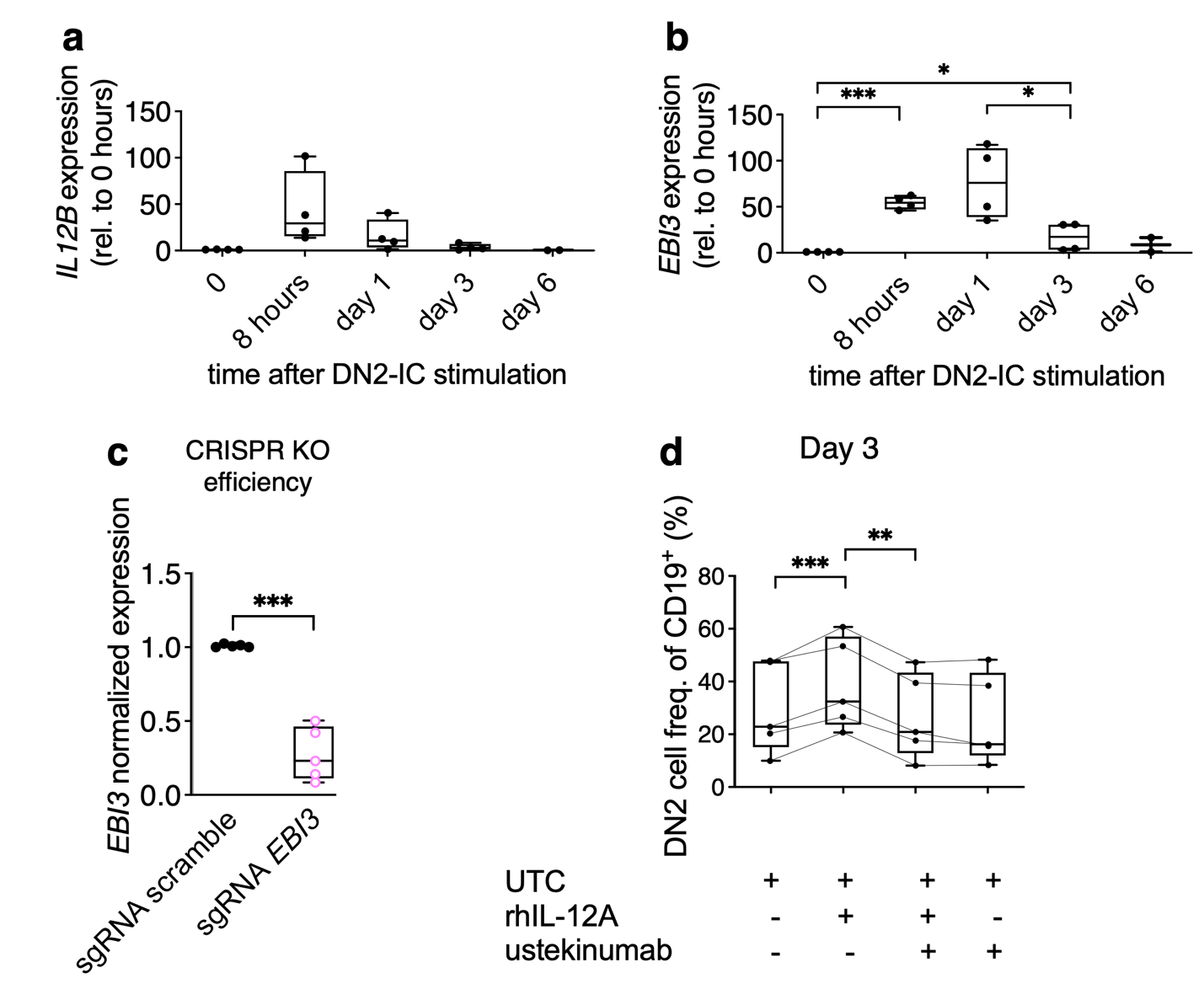

**Extended Figure 2. IL-12A-driven DN2 B cell differentiation is dependent on IL-12.**

**(a-b)** gene expression kinetics of *IL12B* **(a)** and *EBI3* **(b)** in naïve B cells following DN2-IC stimulation at the indicated time points. **(c)** *EBI3* CRISPR KO efficiency, measured by *EBI3* gene expression at day 3 post-stimulation with the DN2-IC. **(d)** Effect of ustekinumab on IL-12A-mediated DN2 B cell differentiation at day 3 post-DN2-IC stimulation. Data are presented as box plots showing the median and 25th/75th percentiles; whiskers indicate the range. Individual data points represent biological replicates (donors, n = 4 to 5). Cell populations are calculated as percentages of total CD19^+^ B cells. scramble sgRNA: cells nucleofected with the scramble sgRNA. sgRNA *EBI3*: cells nucleofected with sgRNA targeting *EBI3* gene. UTC: untreated control, naive B cells stimulated with the DN2-IC. RhIL-12A: naive B cells stimulated with the DN2-IC in the presence of rhIL12A (200ng/ml). Ustekinumab: naive B cells stimulated with the DN2-IC in the presence of ustekinumab (10ug/ml). For all qPCR data involving DN2-IC stimulation across the manuscript, *RPL10A* was utilized as the primary housekeeping control due to its high stability in this model. In panel **a-b**, where *GAPDH* expression fluctuated across the stimulation time-course, data are expressed relative to the Ct values at time 0 (naïve B cells). These expression trends for *IL12B* and *EBI3* were independently validated in separate experiments using *MTHFR* as a stable reference, showing consistent results with the time 0 normalization. In panel **c**, gene expression is normalized to *GAPDH* and expressed relative to the scrambled (scramble sgRNA) control. Two-tailed paired t-test: **p* < 0.05, *p* < 0.01, ****p* < 0.001, *****p* < 0.001.

**Extended Figure 3.**

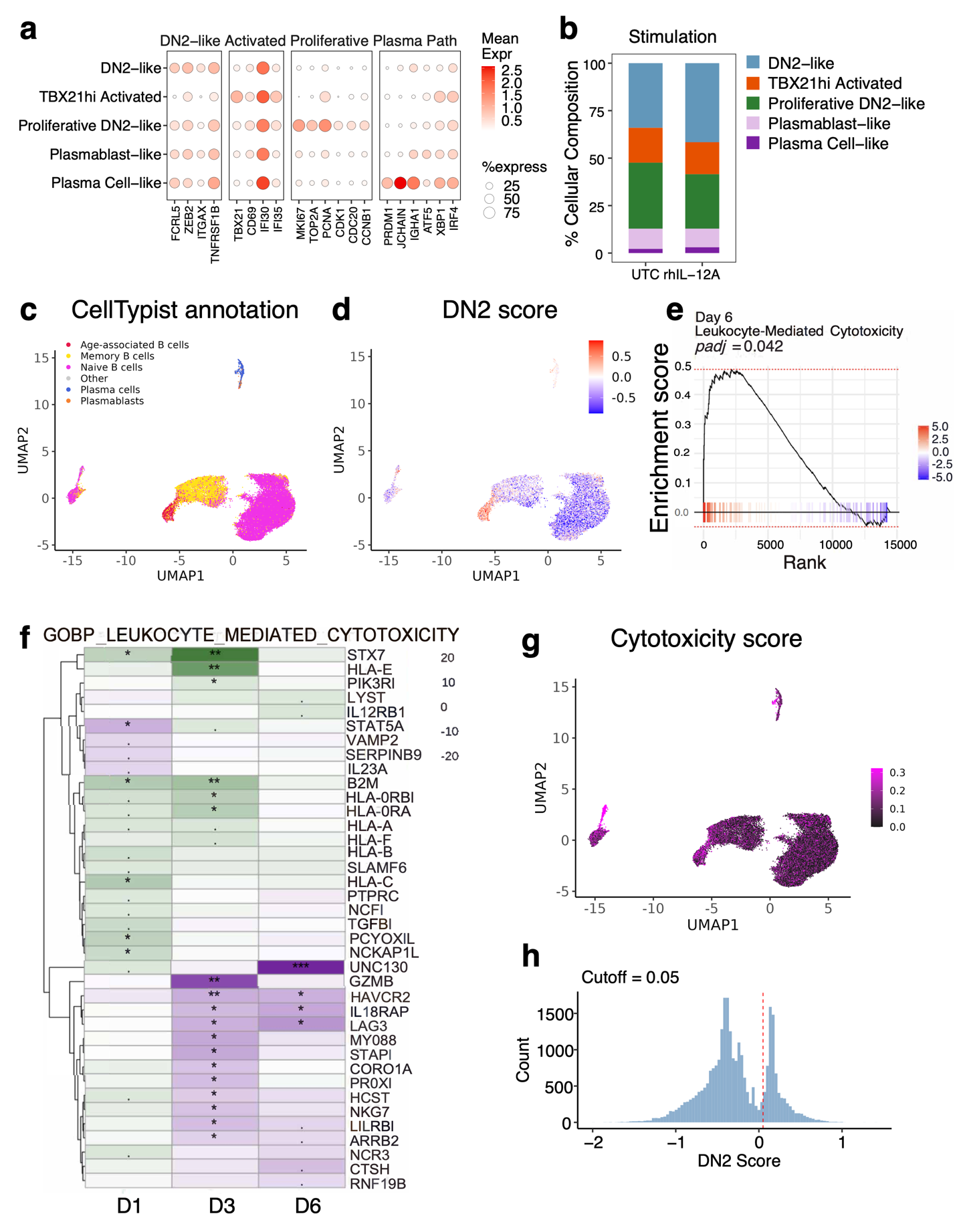

**Extended Figure 3. DN2 B cells inherently possess a previously unrecognized cytotoxic program. (a)** Dot plot showing normalized expression of marker genes used for cell-state identification **(b)** Bar plot showing the percentages of the main cell state within IL-12A stimulation (rhIL-12A) and untreated control (UTC) conditions. **(c)** UMAP visualization showing CellTypist annotation of the B cells from the PBMCs dataset from Nehar-Belaid et al., Nature Immunology (2020). **(d)** UMAP visualization projecting the calculated DN2 score onto the same dataset. **(e-f)** Gene Set Enrichment Analysis (GSEA) **(e)** demonstrating enrichment of the Leukocyte-Mediated Cytotoxicity pathway under the IL-12A condition at day 6, alongside the heatmap **(f)** showing t-values of the genes from the gene set that are expressed across all timepoints and have p<0.01 at one timepoint at least. **(g)** UMAP visualization projecting the calculated cytotoxicity score onto the Nehar-Belaid et al., Nature Immunology (2020) dataset in B cells only. **(h)** Histogram showing bimodal distribution of DN2 score, with cutoff at 0.05. In **(e,f)**, the t-values and p-values were calculated from linear mixed models at each timepoint; *p < 0.05, p < 0.01, ***p < 0.001, or where indicated, FDR-adjusted p-values (padj) are explicitly shown.

**Extended Figure 4.**

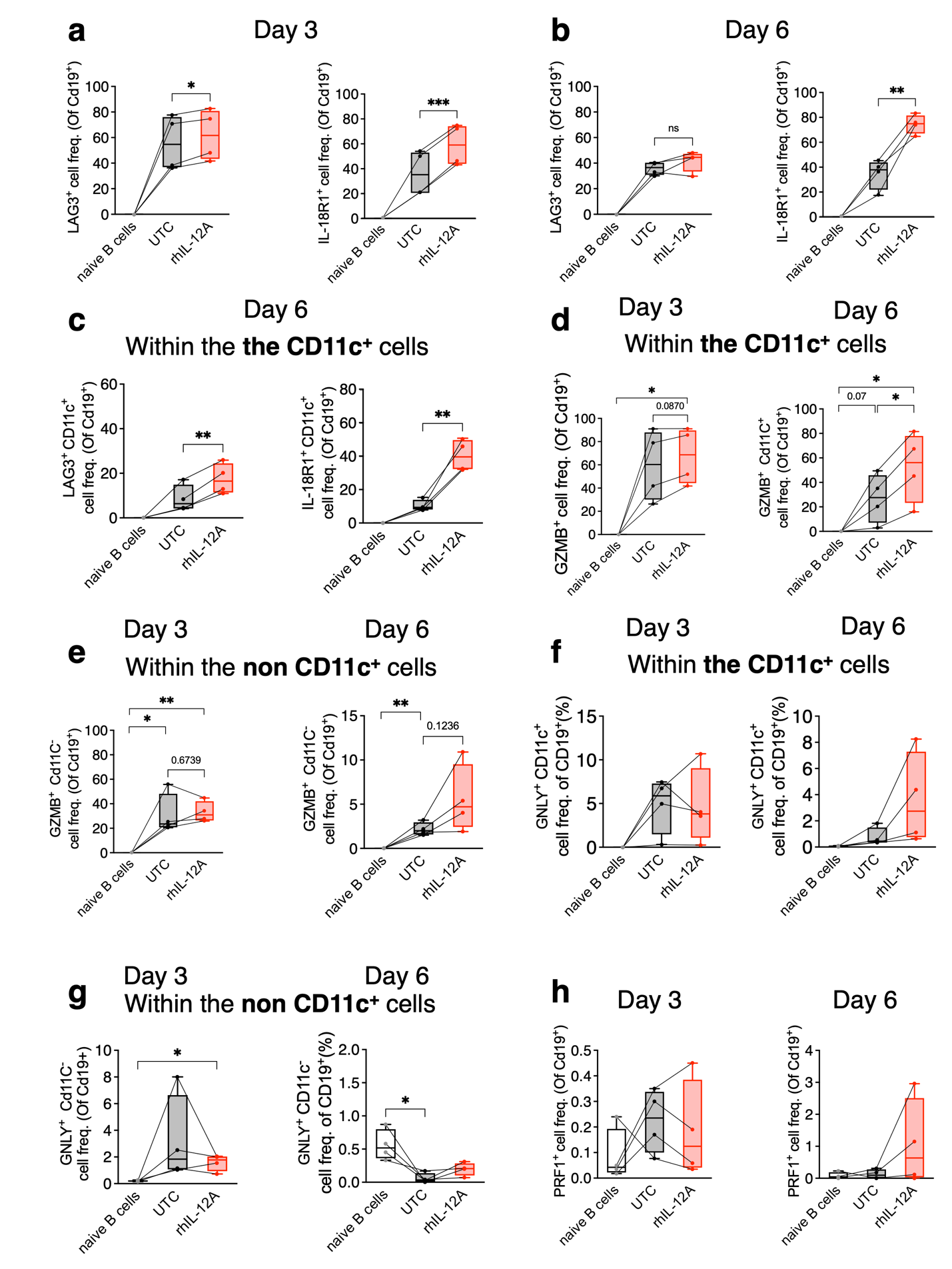

**Extended Figure 4. DN2 B cells cytotoxic program is significantly potentiated by IL-12A signaling.**

**(a-b)** Frequencies of LAG3^+^ and IL-18R1^+^ B cells following naïve B cell stimulation with DN2-IC at day 3 **(a)** and day 6 **(b)**. **(c)** Frequencies of LAG3^+^ and IL-18R1^+^ cells specifically within CD11c^+^ B cell compartment at day 6 post-stimulation. **(d-e)** Percentages of GZMB^+^ cells within the CD11c^+^ **(d)** or the non-CD11c^+^ **(e)** B cell subset at day 3 and day 6 post-stimulation. **(f-g)** Percentages of GNLY^+^ cells within the CD11c^+^ **(f)** or the non-CD11c^+^ **(g)** B cell subset at day 3 and day 6 post-stimulation. **(h)** Percentages of PRF1^+^ B cells at day 3 and day 6 post-stimulation. Data are presented as box plots showing the median and 25th/75th percentiles; whiskers indicate the range. Individual data points represent biological replicates (donors, n = 3 to 4). Naïve B cells: unstimulated naïve B cells at baseline. UTC (untreated control): naïve B cells stimulated with DN2-IC alone. RhIL-12A: naïve B cells stimulated with DN2-IC in the presence of recombinant human IL-12A. Cell populations are calculated as percentages of total CD19^+^ B cells. Two-tailed Paired t-test: **p* < 0.05, ***p* < 0.01, ****p* < 0.001.

**Extended Figure 5.**

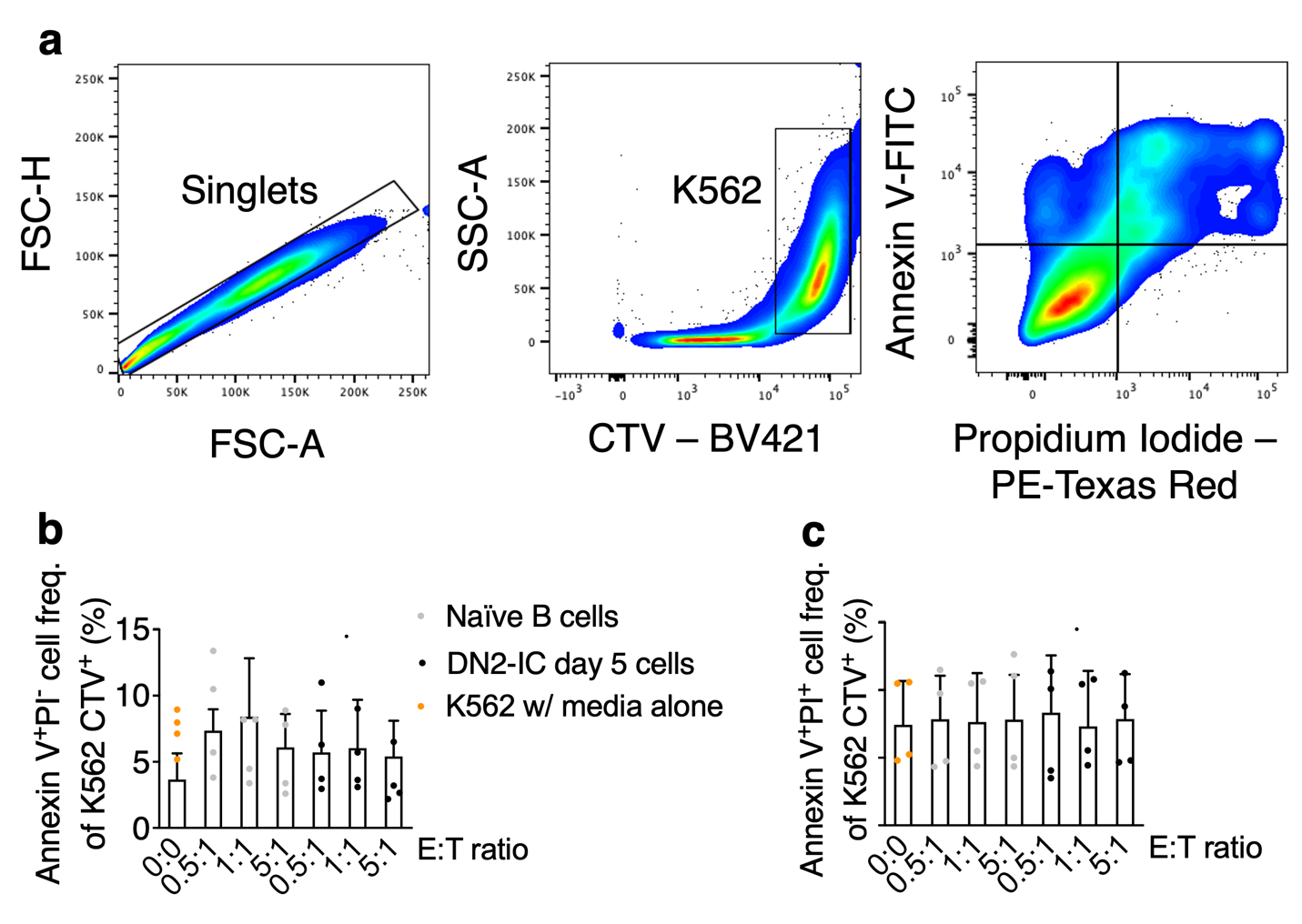

**Extended Figure 5. *In vitro*-generated DN2 B cells can kill K562 cells.**

**(a)** Representative gating strategy for FACS-based cytotoxicity assays using a K562-DN2 cell co-culture system. K562 target cells were labelled with CellTrace Violet (CTV) and co-cultured for 4 hours with *in vitro*-generated DN2 B cells at day 5 post stimulation with the DN2-IC at indicated effector-to-target (E:T) ratios. Naïve B cells stimulated with BAFF overnight were used as negative control. Target cell death was assessed by Annexin V and Propidium Iodide (PI) staining within the K562 CTV^+^ population. **(b, c)** Quantification of Annexin V^+^ PI^+^ (late apoptotic/necrotic) **(b)** and Annexin V^+^ PI^-^ (early apoptotic) **(c)** CTV^+^ K562 cells across various E:T ratios and stimulation conditions. Naïve B cells: naïve B cells stimulated with BAFF (10 ng/mL) overnight (negative control). DN2-IC day 5 cells: naïve B cells differentiated for 5 days with the DN2-IC cocktail. K562 w/ media alone: target cells cultured in the absence of effector B cells to determine baseline spontaneous cell death. Data are presented as mean ± SD. Each dot represents an individual donor (n = 4). Cell populations are calculated as a percentage of CTV^+^ K562 cells.

**Extended Figure 6.**

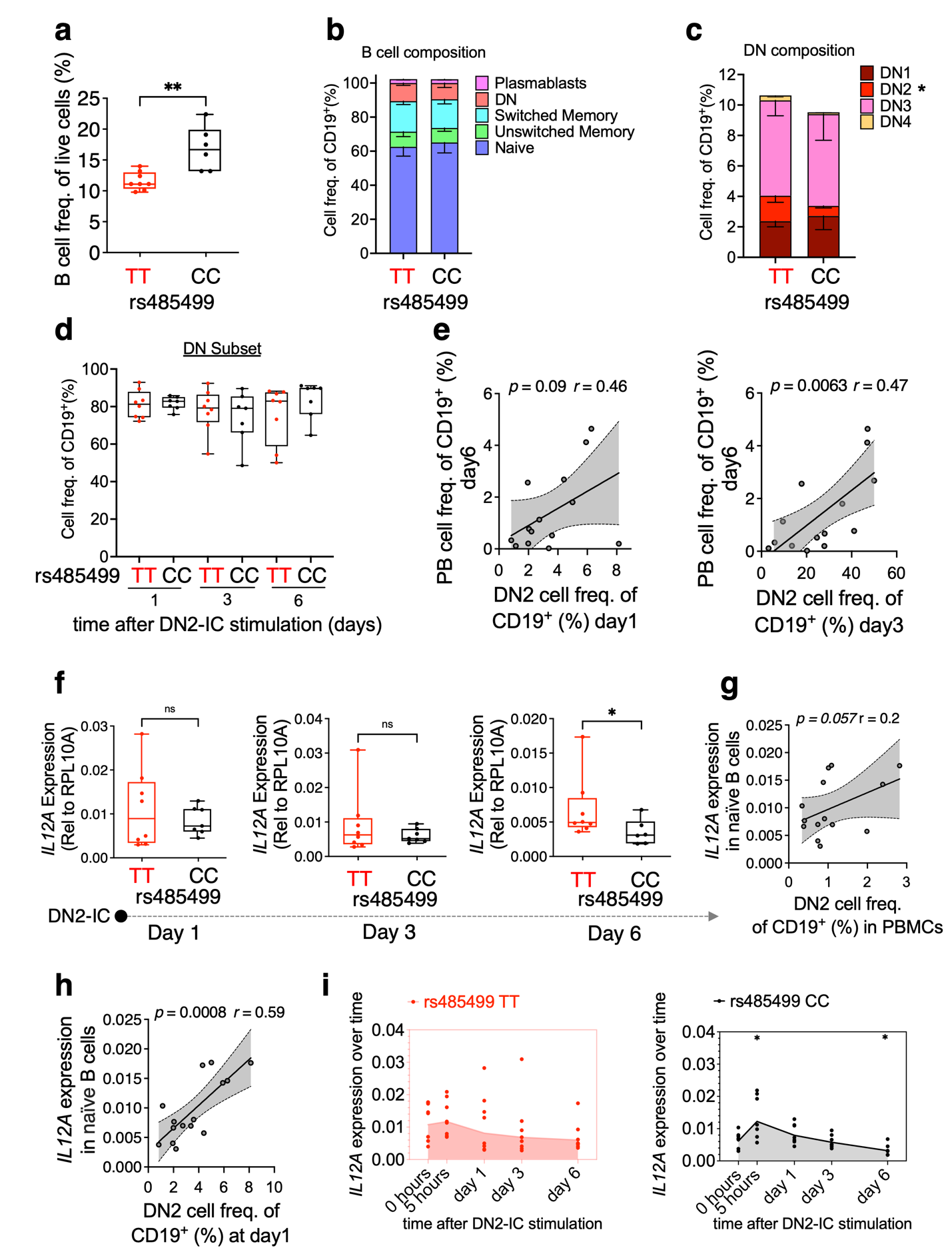

**Extended Figure 6. rs485499 risk allele is associated with DN2 expansion and enhanced B cell *IL12A* production.**

**(a)** Frequency of total CD19^+^ B cells within the live cell population in PBMCs from donors homozygous for the rs485499 risk allele (TT, n = 8) and non-risk allele (CC, n = 7). **(b, c)** Overall B cell composition **(b)** and DN subset distribution **(c)** in PBMCs stratified by rs485499 genotype. In **c** asterisk (*) denotes a statistically significant difference in the DN2 subset (**p* < 0.05, one-tailed Mann-Whitney U test). **(d)** Longitudinal DN composition of naïve B cells from risk and non-risk carriers undergoing *in vitro* differentiation with the DN2-IC at the indicated time points. **(e)** Correlation analysis between the frequency of *in vitro* generated DN2 B cells (at day 1 and day 3) and plasmablast (PB) frequencies at day 6. **(f)** *IL12A* gene expression kinetics were measured at days 1, 3, and 6 post-stimulations with DN2-IC in risk (TT) and non-risk (CC) cohorts. **(g, h)** Correlation between naïve B cell-derived *IL12A* and DN2 expansion in PBMCs **(g)** or at day 1 post-stimulation (**h**). **(i)** *IL12A* expression kinetics in naïve B cells from risk allele carriers (right panel) and non-risk carriers (left panel) stimulated with DN2-IC. Data in panels **(a, d and f)** are presented as box plots showing the median and 25th/75th percentiles; whiskers indicate the range. Individual data points represent biological replicates (donors, n= 7 to 8). Data in panels **(b and c)** are presented as mean ± SEM. Cell populations are calculated as percentages of total CD19^+^ B cells. **(f-h)** Gene expression is normalized to *RPL10A* **(f)** and expressed relative to time 0 of stimulation (naïve B cells) **(f)** using the ΔΔCt method. One-tailed Mann-Whitney U test **p* < 0.05, ***p* < 0.01. For correlation plots **(e, g, h)**, Pearson’s correlation coefficient (*r*) and associated one-tailed p-values are shown. The regression line is shown in black, with the gray shaded area representing the 95% confidence interval.

**Extended Figure 7.**

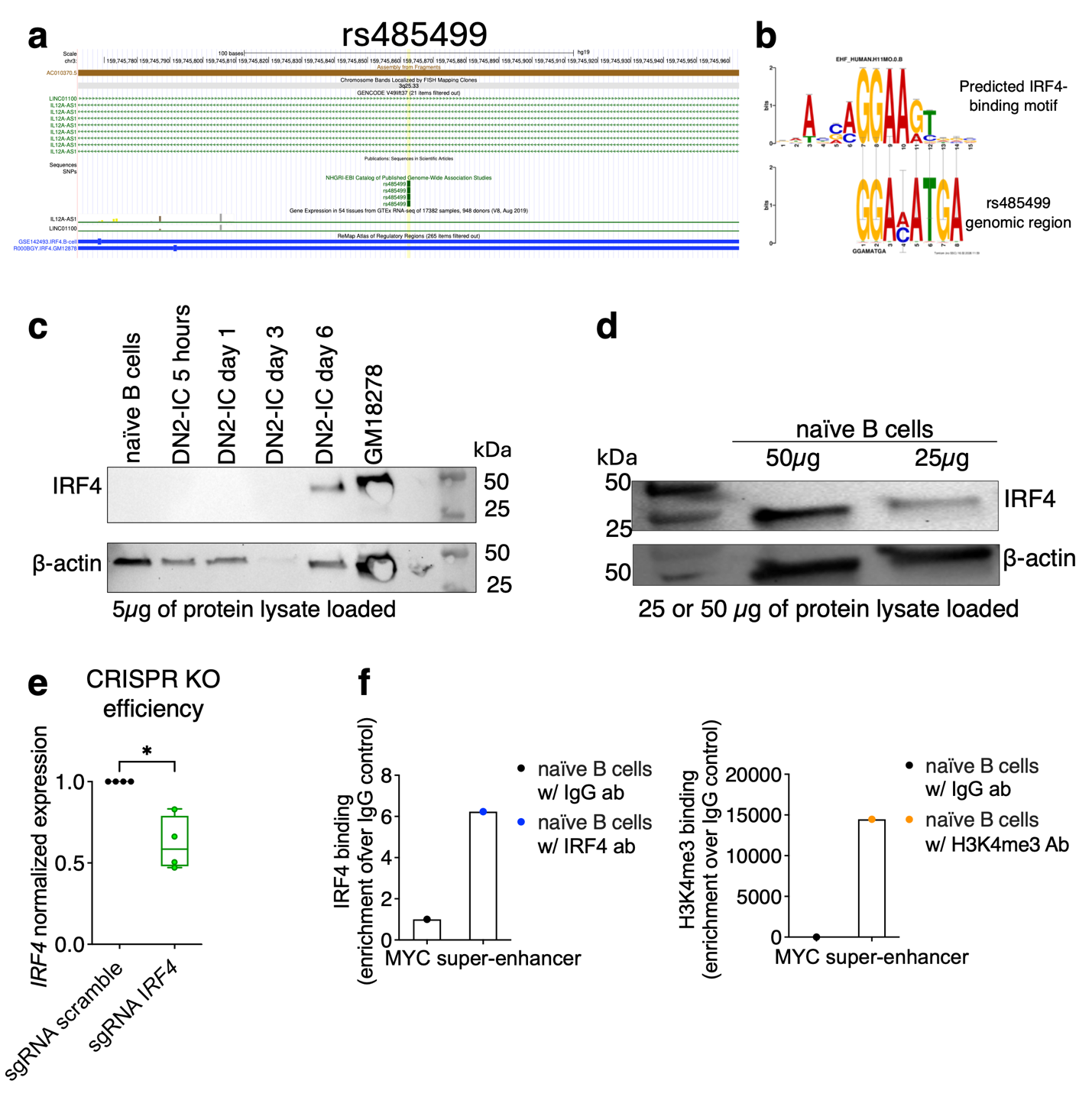

**Extended Figure 7. rs485499 risk allele is associated with enhanced IRF4 binding in naïve B cells.**

**(a)** ChIP-seq tracks demonstrate IRF4 binding peaks at the rs485499 region in the GM12878 cell line (ENCSR000BGY^2^) and in human peripheral blood-derived B cells stimulated with a plasmablast-inducing cocktail (GSE142493^3^). **(b)** Sequence logo comparison generated by MEME and Tomtom analysis, illustrating similarities between the predicted IRF4-binding motif (AGGAAGT) and the rs485499-containing genomic region. **(c, d)** Western blot analysis of IRF4 protein expression kinetics in naïve B cells following DN2-IC stimulation **(c)** and in unstimulated naïve B cells **(d).** β-Actin served as the loading control. The amount of protein loaded is reported in the figures. **(e)** *IRF4* gene expression levels following CRISPR-mediated *IRF4* knockout in naïve B cells stimulated with DN2-IC, measured at days 4–6 post-stimulation. **(f)** CUT&RUN-qPCR analysis showing enrichment of IRF4 (left) and H3K4me3 (right) at the *MYC* super-enhancer region in naïve B cells, serving as a positive control for IRF4 recruitment and CUT&RUN assay. Data in panel **(e)** are presented as box plots showing the median and 25th/75th percentiles; whiskers indicate the range. Individual data points represent biological replicates (donors, n=4). Gene expression is normalized to *RPL10A* and expressed relative to the non-targeting scramble control using the ΔΔCt method. Data from two independent experimental runs were combined and plotted together in **e**. Data in panels **(f)** are normalized to IgG control values using the ΔΔCt method.

**EXTENDED REFERENCE LIST.**

1	Bailey, T. L., Johnson, J., Grant, C. E. & Noble, W. S. The MEME Suite. *Nucleic Acids Res* **43**, W39-49 (2015). <https://doi.org:10.1093/nar/gkv416>

2	Zhang, J. *et al.* An integrative ENCODE resource for cancer genomics. *Nat Commun* **11**, 3696 (2020). <https://doi.org:10.1038/s41467-020-14743-w>

3	Cocco, M. *et al.* A dichotomy of gene regulatory associations during the activated B-cell to plasmablast transition. *Life Sci Alliance* **3** (2020). <https://doi.org:10.26508/lsa.202000654>
